# Genomic and phenotypic correlates of mosaic loss of chromosome Y in blood

**DOI:** 10.1101/2024.04.16.24305851

**Authors:** Yasminka A. Jakubek, Xiaolong Ma, Adrienne M. Stilp, Fulong Yu, Jason Bacon, Justin W. Wong, Francois Aguet, Kristin Ardlie, Donna Arnett, Kathleen Barnes, Joshua C. Bis, Tom Blackwell, Lewis C. Becker, Eric Boerwinkle, Russell P. Bowler, Matthew J. Budoff, April P. Carson, Jiawen Chen, Michael H. Cho, Josef Coresh, Nancy Cox, Paul S. de Vries, Dawn L. DeMeo, David W. Fardo, Myriam Fornage, Xiuqing Guo, Michael E. Hall, Nancy Heard-Costa, Bertha Hidalgo, Marguerite Ryan Irvin, Andrew D. Johnson, Eimear E. Kenny, Dan Levy, Yun Li, Joao AC. Lima, Yongmei Liu, Ruth J.F. Loos, Mitchell J. Machiela, Rasika A. Mathias, Braxton D. Mitchell, Joanne Murabito, Josyf C. Mychaleckyj, Kari North, Peter Orchard, Stephen CJ. Parker, Yash Pershad, Patricia A. Peyser, Katherine A. Pratte, Bruce M. Psaty, Laura M. Raffield, Susan Redline, Stephen S. Rich, Jerome I. Rotter, Sanjiv J. Shah, Jennifer A. Smith, Aaron P. Smith, Albert Smith, Margaret Taub, Hemant K. Tiwari, Russell Tracy, Bjoernar Tuftin, Alexander G. Bick, Vijay G. Sankaran, Alexander P. Reiner, Paul Scheet, Paul L. Auer

## Abstract

Mosaic loss of Y (mLOY) is the most common somatic chromosomal alteration detected in human blood. The presence of mLOY is associated with altered blood cell counts and increased risk of Alzheimer’s disease, solid tumors, and other age-related diseases. We sought to gain a better understanding of genetic drivers and associated phenotypes of mLOY through analyses of whole genome sequencing of a large set of genetically diverse males from the Trans-Omics for Precision Medicine (TOPMed) program. This approach enabled us to identify differences in mLOY frequencies across populations defined by genetic similarity, revealing a higher frequency of mLOY in the European American (EA) ancestry group compared to those of Hispanic American (HA), African American (AA), and East Asian (EAS) ancestry. Further, we identified two genes (*CFHR1* and *LRP6*) that harbor multiple rare, putatively deleterious variants associated with mLOY susceptibility, show that subsets of human hematopoietic stem cells are enriched for activity of mLOY susceptibility variants, and that certain alleles on chromosome Y are more likely to be lost than others.

## Introduction

A cell that has acquired a somatic mutation can undergo clonal expansion which leads to genetic mosaicism, the presence of genetically distinct cell populations in a multicellular organism. Genetic mosaicism is detectable in otherwise healthy tissues, of which blood has been most extensively studied. The most common somatic chromosomal alteration observed in blood is loss of the sex chromosomes, with mosaic loss of Y (mLOY) in males being more common than loss of chromosome X in females. The frequency of mLOY increases non-linearly with age and is detectable in over 30% of males over the age of 70^1^. Varying levels of evidence support associations between mLOY and increased risk of several aging-related diseases including solid cancers, hematologic cancers, cardiovascular disease, and Alzheimer’s disease^2–9^.

To date, mLOY has been studied in cohorts of predominantly European and Japanese ancestry using genome-wide SNP array intensity data^1,10^. In this study we sought to better understand mLOY biology and its clinical and demographic correlates by surveying mLOY in 25,517 ancestrally diverse male participants in the Trans Omics for Precision Medicine (TOPMed) program. In addition to the ancestral diversity in TOPMed, these WGS data provided us the opportunity to compare the sensitivity of WGS-based mLOY calls to those called from arrays, comprehensively investigate the common and rare germ-line drivers of mLOY and its phenotypic correlates, and study the co-occurence of mLOY with other types of somatic mutations.

## Results

We utilized allele-specific read depth of heterozygous markers within the pseudo-autosomal region 1 (PAR1) of the X/Y chromosomes from ∼38X whole genome sequencing (WGS) of leukocyte DNA to detect mLOY in 25,517 ancestrally diverse male participants from 13 different studies in the Trans Omics for Precision Medicine (TOPMed) program. The participants ranged in age from 3 to 95 years (mean = 56; SD = 14.5) and included 7,653 individuals of African American ancestry (AA), 2,838 individuals of Hispanic ancestry (HA), 14,059 individuals of European ancestry (EA), and 501 individuals of East Asian ancestry (EAS) (**Figure 1A**). Mosaic loss of Y (mLOY) was detected from WGS in 2,870 males or 11.5% of 25,051 surveyed. In individuals with detectable mLOY the median estimated mutant cell fraction (CF) was 13.4% (range 4.4% - 100% **Figure 1C**). One-third (33%) of those with mLOY had an estimated CF less than 10%.

**Figure 1.**
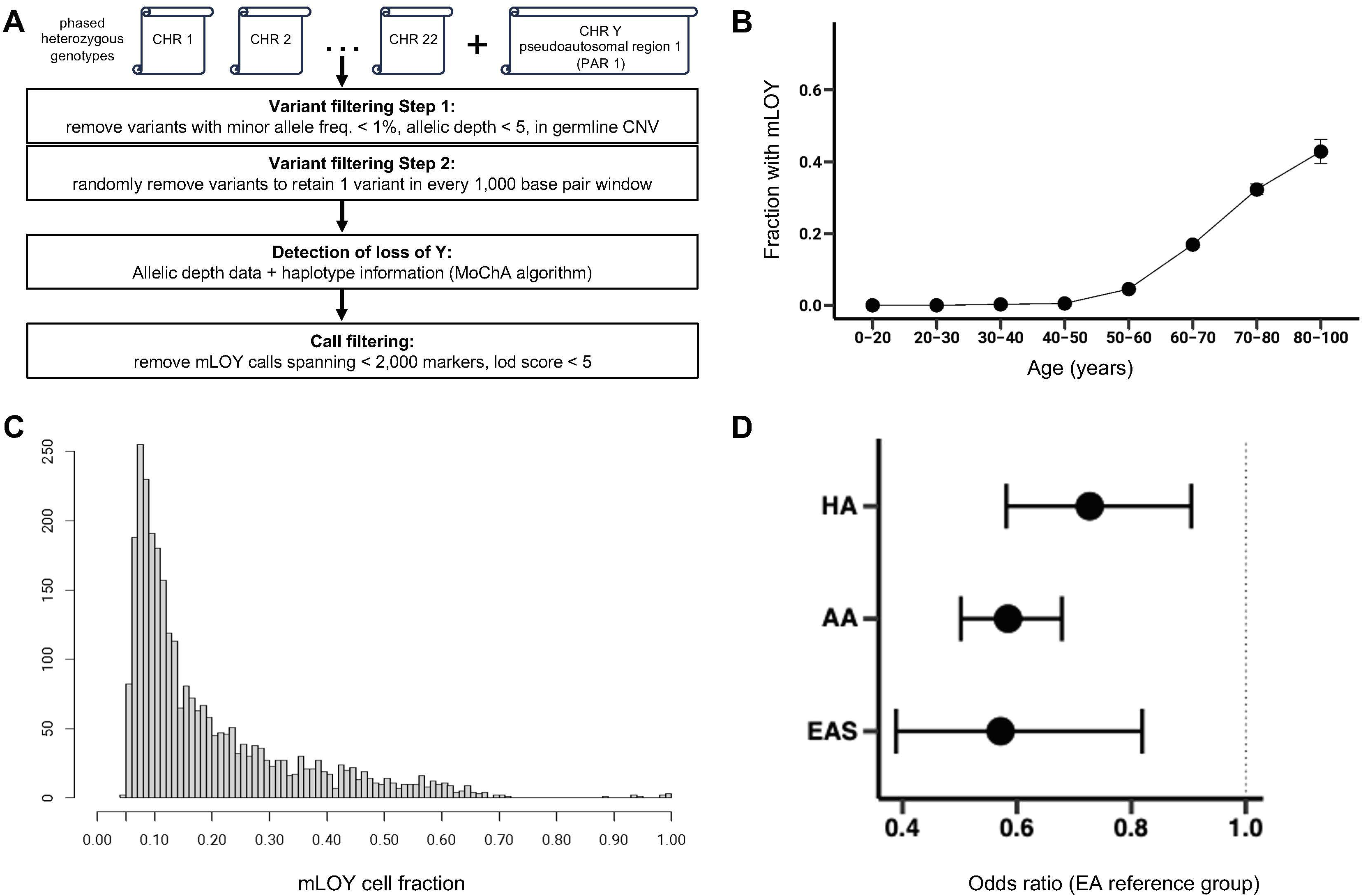
**A)** Overview of data processing for detection of mosaic loss of chromosome Y (mLOY). **B)** Fraction of samples (y-axis) with mLOY by age (years) bins (x-axis). **C)** Histogram of the count of samples (y-axis) and the estimated fraction of cells in the samples with mLOY (x-axis). **D)** mLOY odds ratio (adjusted for covariates) for African American (AA), Hispanic American (HA), and East Asian (EAS) ancestry individuals compared to European American (EA) ancestry individuals.

We compared the mLOY calls from the WGS data with mLOY calls obtained from genotyping arrays on a subset of 2,450 TOPMed males from the MESA study (**Supplementary Materials**). The WGS data were substantially more sensitive, detecting over 5 times the mLOY cases than the genotyping array data, with detection improvements most notable for low cell fraction (CF) mLOY. We also compared the TOPMed haplotype-based mLOY calls to chromosome Y read depth normalized to autosomal read depth, and found that read depth tracked with mLOY status, particularly for mLOY events with CF > 10% (**Supplementary Figure 6**) but that read-depth alone could not effectively recapitulate mLOY status. These results suggest that haplotype-based approaches coupled with WGS data are essential for detecting mLOY events across the spectrum of CFs.

### Association of mLOY with age, smoking, and genetic ancestry

As observed in previous studies, the prevalence of mLOY increased exponentially with age (**Figure 1B**). Among the subset of 14,726 male TOPMed participants for whom smoking status was available, we confirmed the previously reported association between smoking (ever vs. never) and increased risk of mLOY (OR=1.49, P=5.46e-9)^11–13^, and within the group of ever smokers a positive association between pack-years and mLOY (OR=1.0039, P=0.0067).

Individuals were assigned to genetic ancestry groups using the HARE algorithm^14^. Consistent with prior reports^6,10^, (**Tables S1-S2, & Supplementary Figure 1**), we observed lower age-adjusted frequencies of mLOY in AA and EAS compared to EA (AA OR=0.69, P=1.6e-7; EAS OR=0.54, P=6.6e-4). We also found that HA had a lower frequency of mLOY compared to EA (OR=0.80, P=0.048). The overall higher frequencies of mLOY in the EA group remained significant when the analyses were adjusted for smoking status (**Table S2**).

We have previously shown that the power to detect somatic chromosomal events (such as mLOY) is dependent on the observed number of heterozygous sites^15^. The number of heterozygous markers is not uniform across different ancestry groups and thus could bias cross-ancestry comparisons of mLOY rates. Therefore, we down-sampled heterozygous markers in order to achieve equal heterozygosity rates across genetic similarity groups and then re-ran the mLOY detection pipeline in this down-sampled data set (see **Methods**)^15^. The down-sampling procedure resulted in a decrease in the number of mLOY calls, with the largest change observed for the AA group which has the highest heterozygosity rate in PAR1 (**Table S3**). The association between genetic ancestry and mLOY remained significant after the adjustment in heterozygosity rate with all estimated ORs < 0.73 for AA, EAS, and HA compared to EA (**Figure 1D** and **Table S4**). The observed differences remained significant after further adjustment for smoking and the estimated effect size was more pronounced (ORs<0.61 for AA, EAS, and HA compared to EA see **Table S4**). The associations between ancestry and mLOY were consistent for low (<10%) and high (>=10%) mutant cell fraction (CF) mLOY (**Table S5**).

Individuals from the AA group have varying proportions of European and African continental genetic ancestry, allowing us to directly test for an association between proportion of continental ancestry and mLOY. We found that the proportion of European continental ancestry across autosomal chromosomes within the AA group had a positive association with mLOY (P=0.0026) (**Table S6**). Individuals from the HA group have varying levels of African, European, and American continental ancestry. In this group, European continental ancestry was also positively associated with mLOY (P=0.011) (**Table S7**). By contrast, there was no association of mLOY with American continental ancestry among HA (P=0.41) (**Table S7**). We repeated these analyses adjusting for smoking and found that the positive association between European ancestry and mLOY in the AA and HA group were consistent (AA P=0.013; HA P=0.013) (**Tables S6-S7**).

### Genetic associations with mLOY

We tested for associations between germline genetic variants and mLOY, nominally replicating 90 of 248 associations (at P<0.05 with consistent direction of effect) from the largest previously reported mLOY GWAS (**Tables S8**)^16^. Genetic variants at 4 genomic loci reached genome-wide significance (P<5e-8) in TOPMed; two of these were proximal to *TCL1A* and *BCL2L1* (**Table 1**) which have been previously reported. The additional loci near *TSC22D2* and *QK1* may impact gene expression in blood cells as shown by Hi-C expression data for *TSC22D2* and data which implicates *QK1* in erythropoiesis^17,18^. We investigated differences in the allele frequency of the alleles at the 4 genome-wide significant loci across groups of individuals with shared genetic ancestry. Three of the 4 protective alleles are more common in AA compared to EA. Notably, the frequencies of protective alleles at *TCL1A* and *BCL2L1* alleles are approximately twice as common in AA compared to EA (**Table 1**).

**Table 1:**
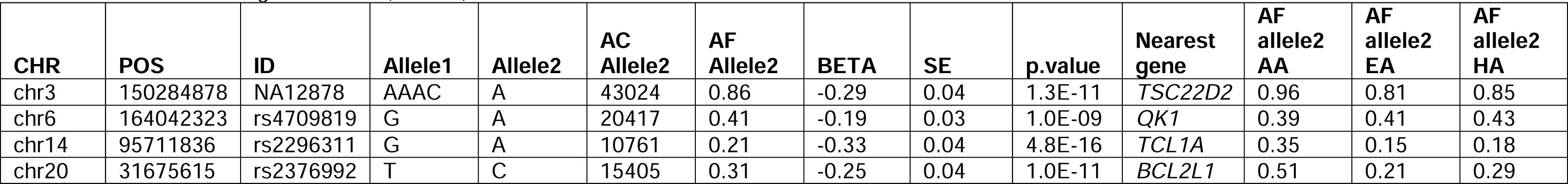
mLOY GWAS significant hits (P<5e-8).

In order to better understand the contribution of the variants in the 4 genome-wide significant loci to differences in mLOY frequencies across ancestry groups, we re-ran the original mLOY-genetic ancestry association analyses of the down-sampled dataset adjusting for genotypes at these loci (**Table S9**). The direction of the associations across ancestry groups were consistent after adjusting for these alleles and the effect size was reduced for AA and HA relative to EA (**Table S9**). In the genetic ancestry analyses conducted among admixed AA, the association of mLOY with European continental ancestry remained significant after adjusting for these variants (P=0.045); though the effect was also attenuated (**Table S6**). For the genetic ancestry analyses conducted among admixed HA, the association of mLOY with European continental ancestry remained significant (P=0.045), but the effect was attenuated (**Table S7**). Additionally adjusting for smoking, the direction of the associations remained consistent (**Table S6-S7**).

In testing for the association between an aggregation of rare-variants (MAF<1%) within each gene and mLOY, we observed significant associations at *CFHR1* (SKAT-O P=2.75e-06) and *LRP6* (SKAT-O P=2.95e-6; **Table S10**). For both genes, the SKAT-O test was driven by a burden of risk increasing alleles (P=8.75e-4 *CFHR1*, P=3.26e-5 *LRP6*) and a mixture of protective and risk increasing alleles (P=3.97e-6 *CFHR1*, P=5.20e-3 *LRP6*). In testing for the contribution of individual rare-variants to these results, we found that multiple putatively loss-of-function variants were driving these associations (**Tables S11-S12**). Interestingly, both of these genes appear highly and relatively selectively expressed in hematopoietic stem cells (HSCs) in comparison with other primary human hematopoietic cell types (**Figure 2**)^19,20^. *LRP6* is required for Wnt/β-catenin signaling which in turn regulates several cellular pathways including cell proliferation and differentiation in hematopoietic stem cells HSCs^21,22^. Finally, we identified 24 genes where rare-variant association with mLOY (P<0.1) were proximal to GWAS associations from Kessler et al. (**Table S13**).

**Figure 2.**
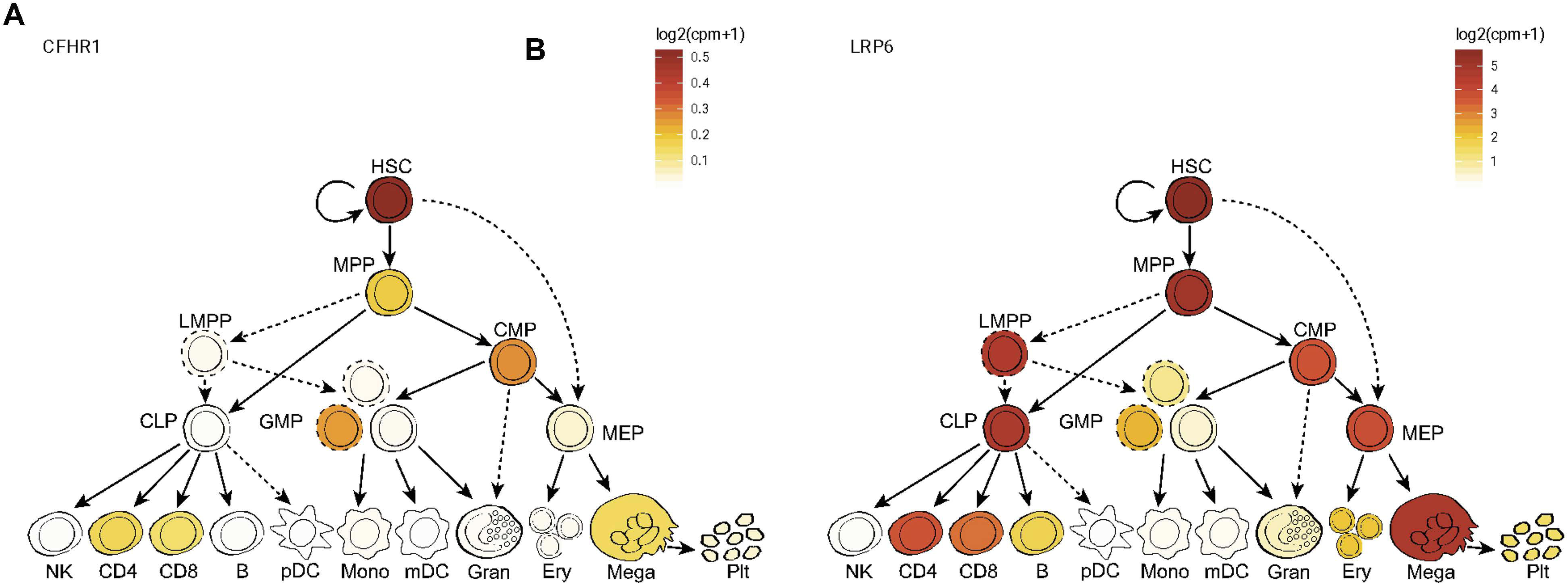
Normalized expression of **A)** CFHR1 and **B)** LRP6 across primary human hematopoietic cell types.

Loss of the Y chromosomes leads to a deviation from the 1-to-1 ratio of alleles at heterozygous sites in PAR1, specifically resulting in a decreased read depth of the allele on Y chromosome relative to the allele on the X chromosome. Under the assumption that alleles on PAR1 are in Hardy-Weinberg equilibrium, both alleles at heterozygous sites should have an equal chance of being on the Y chromosome. To understand whether certain alleles in PAR1 were lost more frequently than others, we performed an allelic shift analysis in males with mLOY and found 59 intronic SNPs in the *XG* and *CD99* genes which show a bias towards one of the two alleles (**Table S14**). We conducted the same analysis in a control set of males without mLOY and did not find statistically significant SNPs. The *XG* gene spans the PAR1 boundary, with exons present in the PAR1 region and other exons exclusive to the X chromosome^23,24^. The *CD99* gene is exclusive to the PAR1 region^23,24^. The rs311103 variant in *XG* defines the XG blood group system^23,24^ but none of the alleles that demonstrated an allelic shift (**Table S14**) were in linkage disequilibrium (LD) with rs311103, suggesting that the allelic shift impacts *CD99* rather than *XG*. Seventeen of the 59 statistically significant SNPs from the allelic shift analyses are expression quantitative trait loci (eQTL) for *CD99* in the TOPMed data set. Of these, 7 are not in LD with each other (LD rsq < 0.1) and the allele associated with increased *CD99* expression was preferentially retained (**Table S15**).

### Co-occurrence between LOY and other types of somatic variation

Previous studies have profiled aging-related genomic changes for the TOPMed datasets included in this study. These data allowed us to assess the co-occurence of mLOY with other types of somatic mutations. We observed that the prevalence of mosaic chromosomal alterations (mCAs) in the autosomes was higher among mLOY carriers (8.7%) than non-carriers (4.1%, OR=2.2 **Table S16**) likely due to the very strong age-dependence of all types of somatic mutations. When adjusted for age, age^2^, genetic ancestry group, and study, there was no overall association between mLOY and presence of an autosomal mCA (OR=1.02; 95% CI 0.87 - 1.20; P=0.78). **Supplementary Figure 2A** shows the frequencies of six common autosomal mCAs by mLOY carrier status. When each of the most common autosomal mCA (1p, 11q, 12p, 12q, 14q, 20q) were assessed individually, 11q was associated with higher prevalence of LOY (OR=2.34; P=0.01), while 20q was associated with lower prevalence of mLOY (OR=0.48, P=0.04) when adjusted for age, age^2^, ancestry, and study. Next, we tested for association between mLOY and mCAs categorized based on their occurrence in hematological malignancies. To do so mCAs were labeled as lymphoid-mCA (L-mCA), myeloid-mCA (M-mCA), ambiguous-mCA (A-mCA) which are common to both lymphoid and myeloid disorders, and unclassified (U-mCAs) which are not associated with either malignancy^25^. Compared to those without an mCA, individuals with U-mCAs were less likely to have mLOY (OR=0.79, P=0.048). Individuals with L-mCAs and A-mCAs were more likely to have mLOY (L-mCAs OR=1.33, P=0.06; A-mCA OR=1.52, P=0.05); however, this difference did not reach statistical significance. We did not observe an association between M-mCAs and mLOY (OR=1.02, P=0.92) (**Table S17**).

Overall, 1,137 or 5.3% of the TOPMed males were carriers of mutations that drive clonal hematopoiesis of indeterminate potential (CHIP), as ascertained in TOPMed (mean VAF=21%, range 6% - 97%). The prevalence of CHIP was ∼2-fold higher in mLOY carriers (9.2%) than non-carriers (4.8%) (OR=2.02, **Table S18**). Yet when adjusted for age, age^2^, ancestry, and study, the presence of CHIP was associated with a *lower* odds of mLOY (OR=0.78; 95%CI 0.66 - 0.92; P=0.004); **Supplementary Figure 2B** shows the frequencies of common CHIP mutations by mLOY carrier status. In particular, the presence of either *DNMT3A* CHIP and multi-gene CHIP subtypes was lower among mLOY carriers (**Table 2**). The association between any CHIP or *DNMT3A* CHIP and lower odds of mLOY did not differ for low (<10%) and high (>=10%) mutant cell fraction (CF) mLOY; for multi-gene CHIP subtype, the association was somewhat stronger for larger mLOY clones than smaller clones.

**Table 2:** The relationship between CHIP gene and LoY carrier status.

| CHIP type | OR | P |
| --- | --- | --- |
| ASXL1 | 0.836 | 0.479 |
| DNMT3A | 0.642 | 0.001 |
| JAK2 | 2.297 | 0.744 |
| Multiple | 0.479 | 0.002 |
| Other | 1.407 | 0.193 |
| Splice | 0.663 | 0.177 |
| TET2 | 1.014 | 0.945 |
| TP53 | 1.865 | 0.229 |
| PPM1D | 1.282 | 0.479 |

When mLOY clone size was modeled as a continuous variable among the subset of 2,481 mLOY carriers and adjusted for age, age^2^, and study, the presence of CHIP was associated with 3.9% higher mLOY clonal fraction (SE=1%) compared to non-CHIP carriers (P<0.0001). Conversely, among 1,137 CHIP+ men, the presence of mLOY was strongly associated with 3.8% lower CHIP clone size (VAF) (P<0.0001). These results suggest that the mLOY clones may outcompete CHIP clones, limiting their expansion.

### Relationship of mLOY to Telomere-length in TOPMed

Adjusting for age, age^2^, ancestry, and study, mLOY was strongly associated with shorter batch-corrected telomere length (TL; P<0.0001) in TOPMed. When modeling mLOY as the outcome, we estimated that each kilobase (kb) of TL is associated with a 27% decrease in the odds of mLOY (OR=0.73). Moreover, among the 2,870 mLOY carriers, larger clonal fraction (modeled as a continuous variable) was associated with 0.015 kb shorter TL (P=0.006). This finding is consistent with reports that genetically predicted longer telomere length is negatively associated with mLOY in UK Biobank individuals^26^.

### Circulating blood-cell counts and mLOY

When adjusted for age at phenotype measurement, age at TOPMed blood draw, ancestry, and study-sequencing phase of TOPMed, mLOY was significantly associated with: lower RBC count (P=0.036); higher MCV (P=0.005); lower MCHC (P=0.029); higher WBC, neutrophil, and monocyte count (all P<0.001); and higher platelet count (P=1.7e-05) (**Table 3**). These results are largely consistent with those previously reported^10,27^. As with CHIP and autosomal mCAs, mLOY is known to increase risk for myelodysplastic syndrome, myeloproliferative neoplasms and leukemia^28,29^. Such individuals, or those with extreme CBC values, were excluded from analysis. Compared to mLOY, the presence of CHIP was associated with a notably different pattern of hematologic traits: lower hemoglobin/hematocrit and higher RDW, but there was no association with WBC or platelet counts^30^. Additional adjustment for CHIP carrier status did not alter any of the mLOY-blood cell trait associations (**Table S19**). Nor was there evidence of effect modification by CHIP of the mLOY-blood cell trait associations.

**Table 3:** Association between blood-cell traits and mLOY.

| trait | Est | SE | pval | n |
| --- | --- | --- | --- | --- |
| BASO | -0.0375255 | 0.0782905 | 0.6317175 | 10475 |
| EOSIN | -0.2442416 | 0.1954689 | 0.2114764 | 10538 |
| HCT | 0.0529366 | 0.1092809 | 0.6281035 | 13706 |
| HGB | -0.0323303 | 0.0371538 | 0.3842201 | 13709 |
| LYMPHS | -0.0101386 | 0.0052007 | 0.0512661 | 11516 |
| MCH | 0.1047961 | 0.0686363 | 0.1268294 | 11987 |
| MCHC | -0.0598984 | 0.0274777 | 0.0292826 | 13545 |
| MCV | 0.479098 | 0.1723566 | 0.0054492 | 12559 |
| MONOS | 0.0255048 | 0.0055464 | 0.0000043 | 11455 |
| MPV | 0.0546213 | 0.0530178 | 0.3029413 | 5425 |
| NEUTRO | 0.035269 | 0.0062563 | 0 | 10629 |
| PLT | 6.5605195 | 1.747338 | 0.0001744 | 13660 |
| RBC | -0.0327767 | 0.0156412 | 0.0361456 | 11660 |
| RDW | -0.028058 | 0.060167 | 0.6409905 | 6584 |
| WBC | 0.0150052 | 0.0034604 | 0.0000146 | 13497 |

Next we investigated associations between blood cell counts separately for low CF (< 10%) and high CF (>=10%) mLOY (**Table S20**). High CF mLOY was associated with higher MCH and MCV and with lower RBC count (P<1e-4). By contrast, low CF mLOY was associated with lower MCH and MCV (P=0.04 for MCH and P=0.07 for MCV), and with higher RBC count (P=0.07). The difference in the effect of low vs high mLOY on MCH, MCV, and RBC were statistically significant (P<0.001). Both high and low CF mLOY were significantly associated with higher monocyte, neutrophil, and white blood cell counts (**Table S20**), whereas high CF mLOY was selectively associated with lower lymphocyte count. Platelet count was also significantly higher among high CF mLOY carriers and also among low CF mLOY carriers, although it did not reach statistical significance in the latter group. The observed associations of low and high CF mLOY remained consistent when the analyses were adjusted for CHIP carrier status (**Table S21**).

Given the strong relationship of mLOY with peripheral blood counts and the shared genetic architecture between these traits, we assessed relevant blood cell types or states underlying the inherited risk loci for mLOY. We employed the SCAVENGE (Single Cell Analysis of Variant Enrichment through Network propagation of GEnomic data) method to elucidate relevant trait-cell state associations^31^. This method overlaps fine-mapping posterior probabilities derived from GWAS of mLOY susceptibility with accessible chromatin enrichments at single-cell resolution by employing a network propagation approach (see **Methods**). Given that our current understanding is that clonal hematopoiesis events like mLOY arise in the HSCs compartment, we observed strong enrichment for mLOY loci in HSCs and other early progenitors, including erythroid and basophilic progenitor populations (**Figure 3A, Supplementary Figure 4A-C**). Given the known heterogeneity within the HSC compartment in humans^32^, we analyzed variation in enriched or depleted cells based on the SCAVENGE trait relevance score (TRS). Remarkably, we noticed that HSCs with stronger enrichment of GATA, RUNX, and HOX motifs, which are thought to underlie a more primitive group of HSCs, had stronger enrichment for the mLOY TRS, while cells with more myeloid bias, as indicated by CEBP and IRF motifs, were depleted in mLOY enrichments (**Figure 3B-E, Supplementary Figure 4E, E**). This illuminates how distinct subsets of HSCs in humans appear to show enrichment for activity of genetic variants associated with mLOY.

**Figure 3.**
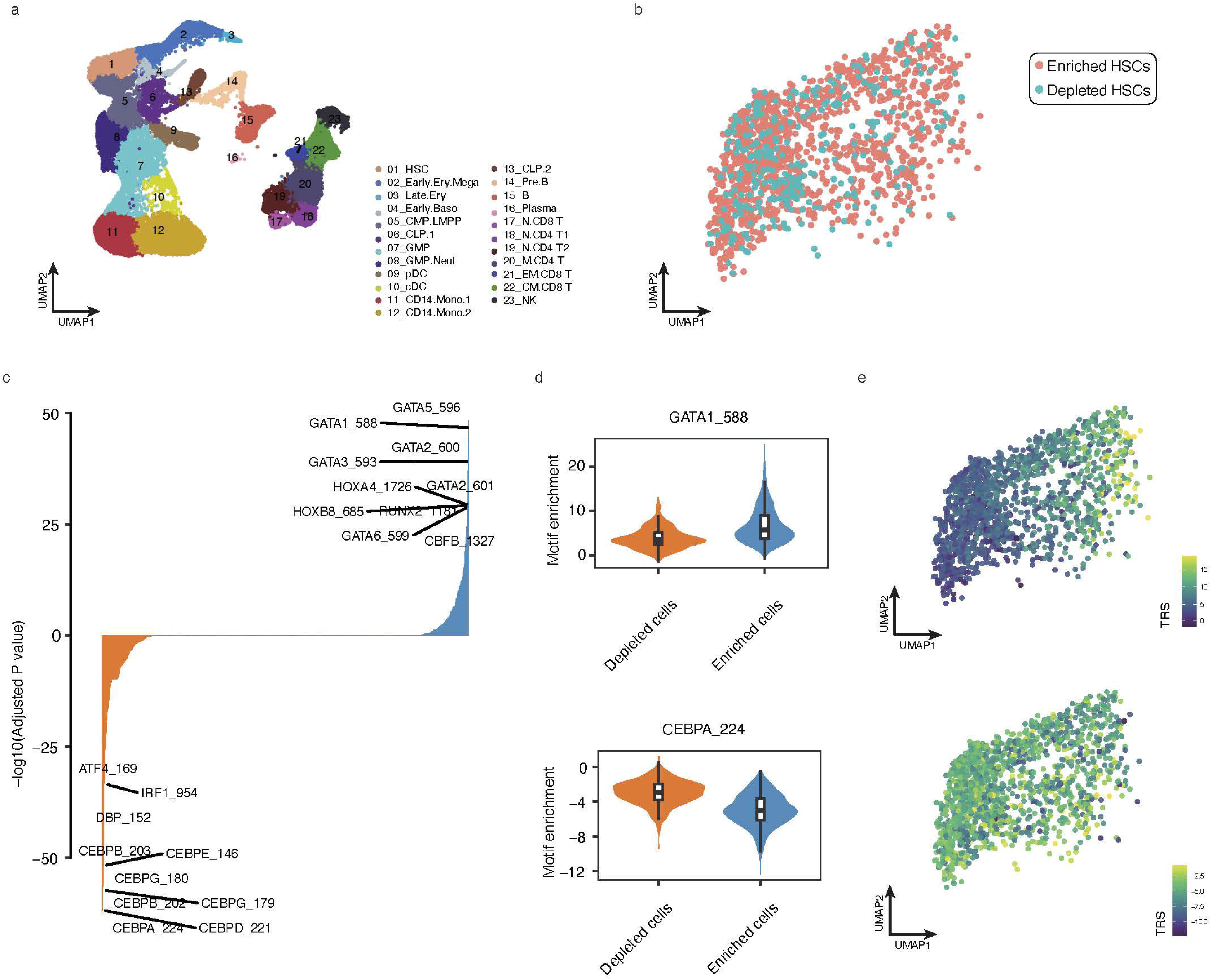
SCAVENGE analysis of the LOY trait using a hematopoietic scATAC-seq dataset. **A)** The uniform manifold approximation and projection (UMAP) plot depicting cell type annotations of scATAC-seq dataset. **B)** The UMAP of hematopoietic stem cells (HSCs) labeled as ‘enriched’ or ‘depleted’, respectively, representing their relevance or non-relevance to mLOY risk by using the permutation tests coupled with SCAVENGE analysis (see methods for details). **C)** Differential comparison of chromVAR TF motif enrichment between LOY risk variant-enriched and variant-depleted HSCs. Bonferroni-adjusted P value is indicated (y-axis) and the TFs were ranked accordingly (x-axis). The selected TF motifs with the top 10 ranks in each direction were labeled. The chromVAR enrichment Z-scores for **D & E)** CEBPA and GATA1 motifs are shown in UMAP plots (right) and violin plots (left) across HSCs as shown in B.

### Gene expression and mLOY

We tested for differentially expressed chromosome Y genes between blood samples from individuals with and without mLOY utilizing expression data available for 2 studies (JHS and MESA) adjusting for smoking as a covariate (see **Methods**). The genes *SLC25A6* and *TMSB4Y* were downregulated in the group with mLOY (adj. P<0.05), as were *CSF2RA*, *RPS4Y1*, and *CD99* (adj. P<0.1) **(Table 4**). Downregulation of *CD99* and *SLC25A6* has been observed in blood, fat, and muscle tissue from individuals with Turner syndrome who are born with a single sex chromosome (chr X) ^33^ and leukocytes with loss of Y have reduced cell surface immunoprotein *CD99* ^34^.

**Table 4:**
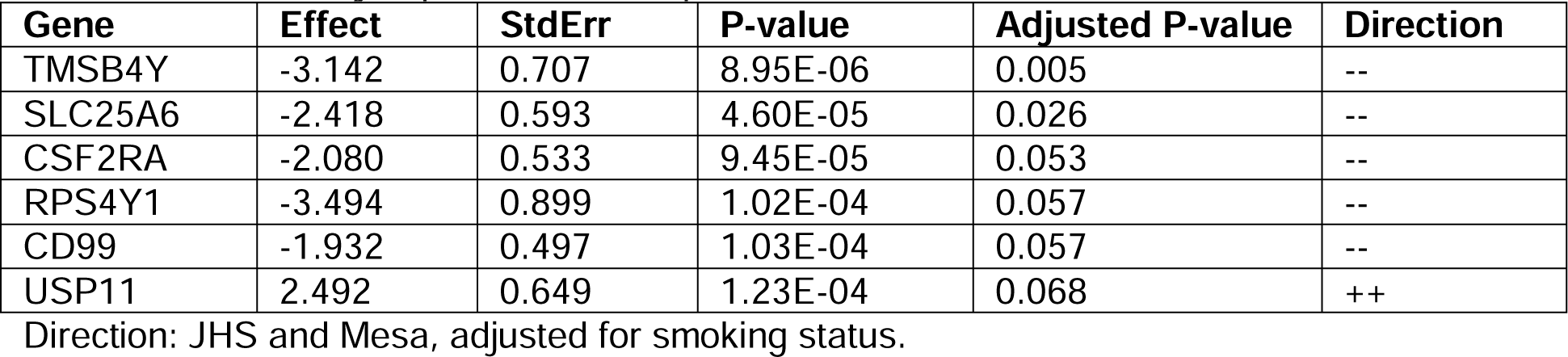
Differentially expressed transcripts between individuals with and without mLOY.

## Discussion

Here, we profiled mLOY in an ancestrally diverse sample of males from the United States with WGS data from the TOPMed project. We used the haplotype-based algorithm (MOsaic CHromosomal Alterations (MoCha)) that relies on well-phased heterozygous markers in the PAR1 of chromosome X to identify mLOY. Mosaic LOY calling using WGS data was more powerful than using array-based genotype data, particularly for mLOY with low clonal fraction. As in a previous study^15^, we found that detection sensitivity for chromosomal alterations such as mLOY is partially determined by the density of heterozygous markers in the region of interest. Accordingly, the relative gain in power with WGS data is dependent on the number of markers on an array in the PAR1 region. For instance, using the TOPMed WGS data we observed a median of 1,022 heterozygous markers in PAR1, compared to a median of 93 heterozygous markers on the Affymetrix 6.0 array. Prior studies of mLOY have used data from the UKBiobank array where they observed an average of 321 heterozygous variants in PAR1 that passed QC^1^. Though read-depth data on chromosome Y from WGS may be used to identify high CF mLOY events (**Supplementary Materials**), the implementation of haplotype-based callers, such as MoCha, are essential for identifying lower CF events.

Our investigation of mLOY in a multi-ancestry cohort confirms the association of mLOY with age, smoking, and genetic ancestry^6,10^. In addition to the previous findings of higher age-adjusted frequencies of mLOY in EA individuals compared to EAS and AA, we also report that EA individuals have higher mLOY frequencies compared to HA individuals. These differences appear to be driven in part by common protective variants at *TCL1A* and *BCL2L1* which are more common in AA and HA. Within each of the admixed AA and HA groups, continental European genetic ancestry was associated with higher prevalence of mLOY when adjusting for the *TCL1A* and *BCL2L1* lead SNPs. Additional, genetic, epigenetic, and environmental factors likely contribute to the increased mLOY frequency in the EA group.

Of mosaic chromosomal alterations, mLOY is the most common and the one with the highest estimated heritability at 32%^1^. In our study, we confirm several mLOY genetic associations including the well-established signal at *TCL1A^1,2,10,11,35^*. The TOPMed WGS data permitted us to conduct aggregate rare-variant association tests with mLOY. We report the association between a burden of rare variants in two genes (*CFHR1* and *LRP6*) with the presence of mLOY. In both cases, putatively loss-of-function variants appear to increase the risk of developing mLOY in blood and both genes appear to be highly and selectively expressed in HSCs in comparison with other primary human hematopoietic cell types. These results suggest that LOY in HSCs may have a proliferative advantage whereby LOY is maintained as progenitor cells differentiate down the blood-cell lineages.

The observed strong association of mLOY with myeloid traits (higher WBC, neutrophil, and monocyte counts), higher platelet count and weaker associations with higher MCV, lower RBC count, and lower lymphocyte count (for high CF mLOY>10%) are largely consistent with findings previously reported in individuals of predominantly European and Japanese ancestry^10,27^. These phenotypic associations may reflect in part shared genetic architecture between mLOY and blood cell traits^27,36^ or shared etiology between mLOY and other clonal hematologic disorders characterized by ineffective erythropoiesis such as myeloproliferative neoplasms^36^ or myelodysplastic syndromes/myeloid leukemias^28,29^.

The observed associations of mLOY with multiple blood cell lineages is consistent with evidence from heritability enrichment analyses suggesting that germline genetic variants associated with mLOY primarily impact clonal expansion of HSC and other early hematopoietic progenitor cells^10^. Our analyses utilizing single cell ATAC-seq data further demonstrates that mLOY polygenicity may be confined to a subset of more primitive HSCs enriched in GATA, RUNX, and HOX motifs, which are also critical hematopoietic transcription factors that control HSC self-renewal and differentiation into diverse mature blood cells. The distinct pattern of blood cell phenotypic associations with CHIP, along with the absence of genetic correlation between CHIP and mLOY^36^ suggests distinct pathogenesis for these two common types of clonal hematopoiesis, which requires further study.

Differential expression analyses confirmed the previously reported down-regulation of *CD99* in mLOY leukocytes^34^. The observation that specific alleles near *CD99* are preferentially lost suggests there is a potential selection for blood cells with reduced *CD99* expression. Differential expression analyses also indicate that *CSF2RA*, which promotes secretion of proinflammatory cytokines as well as HSCs cell proliferation and differentiation^37^, is downregulated in mLOY cells. Future studies which contrast the effect of CHIP and mLOY on expression and inflammatory pathways may help elucidate differences on the impact of these different types of somatic mutations on blood cell traits and disease susceptibility.

Although mLOY and CHIP increase with age, our age-adjusted analyses reveal an overall inverse association between these two types of somatic mutations in blood as well as smaller CHIP clone size among those with mLOY compared to those without mLOY. These results are consistent with a large-scale analysis from the UKBiobank showing an inverse association between age- and smoking-adjusted frequencies of mLOY and CHIP with frequently observed driver mutations such as *DNMT3A* and *TET2*^36,38^. In our CHIP subtype analysis from TOPMed, the inverse phenotypic mLOY-CHIP association was mainly confined to males with *DNMT3A* or multiple CHIP gene drivers. The latter finding is further evidence of an “evolutionary trade-off” between CHIP and mLOY in which competing clonal mutations may limit each other’s expansion frequencies^36,38^. However, individuals with mLOY clones present in greater than 30%-40% of sample cells^39^ have significantly higher odds of carrying CHIP mutations which drive elevated *TCL1A* expression. *DNMT3A* CHIP, which does not drive elevated *TCL1A* expression, is not associated with increased mLOY clone size^35^. Single-cell analyses have demonstrated that mLOY is associated with increased levels of *TCL1A* expression^1^. These results point to a potential synergistic effect of mLOY and certain CHIP mutations on increased *TCL1A* expression and expansion of clones harboring both mutations. Further studies are needed to elucidate the clonal dynamics when independent clones harbor somatic mutations which both drive increased *TCL1A* expression as it is possible that competing clones may limit each other’s expansion. The latter may explain the negative association observed between low mutant cell fraction mLOY and CHIP. The strong association between both mLOY and CHIP with shorter telomere length may also play a role^38^. Higher clonal fraction leading to shorter telomere length may promote genomic instability and acquisition of additional CHIP driver mutations^36,38^. Intriguingly, mLOY and CHIP have opposite associations with Alzheimer’s disease (AD) with mLOY being associated with an ∼2.5 fold increased risk and CHIP having a protective effect^4,40,41^. Additional studies of the clonal dynamics of clones harboring mLOY and CHIP mutations may help further elucidate their association with AD.

## Methods

### Study population

There was a total of 25,051 participants from 13 TOPMed studies (**Table S22**): Genetics of Cardiometabolic Health in the Amish (Amish)^42^, Atherosclerosis Risk in Communities Study (ARIC) ^43^, Barbados Genetics Asthma Study (BAGS), Mount Sinai BioMe Biobank (BioMe)^44^, Coronary Artery Risk Development in Young Adults (CARDIA)^45^, Cleveland Family Study (CFS), Cardiovascular Health Study (CHS)^46^, Genetic Epidemiology of COPD Study (COPDGene)^47^, Framingham Heart Study (FHS)^48^, Genetic Studies of Atherosclerosis Risk (GeneSTAR)^49^, Genetic Epidemiology Network of Arteriopathy (GENOA), Genetics of Lipid Lowering Drugs and Diet Network (GOLDN), Hispanic Community Health Study - Study of Latinos (HCHS_SOL)^50^, Hypertension Genetic Epidemiology Network (HyperGEN), Jackson Heart Study (JHS)^51^, Multi-Ethnic Study of Atherosclerosis (MESA)^52^, and the Vanderbilt BioVU study of African Americans (VU_AF). The HARE machine learning algorithm was used to assign individuals to genetic ancestry subgroups^14^. All study participants provided informed consent and all studies have appropriate institutional review board approval.

### Whole genome sequencing

Data utilized in this study were generated as part of the NHLBI TOPMed program. WGS of blood samples was performed at an average depth of 38X using Illumina X10 technology. Six sequencing centers performed WGS (Broad Genomics, Northwest Genome Institute, Illumina, New York Genome Center, Baylor, and McDonnell Genome Institute). Data used in these analyzes are from ‘Freeze 8’ with reads aligned to human-genome build GRCh38. All centers utilized a shared pipeline for alignment, variant quality control (QC), and sample QC. Variants with excess heterozygosity and mendelian inconsistencies were removed. Sample QC included verification of consistent genetic and annotated sex, concordance between genotypes from WGS and those derived from previous performed array genotyping, and pedigree consistency. More detailed information regarding WGS data processing is provided in Taliun et al.^53^

### Detection of mLOY

Detection of mLOY was performed using allele-specific read depth data from WGS. Mosaic LOY calls were generated using the MOsaic CHromosomal Alterations (MoChA v1.15) software. Input data consisted of read depth at heterozygous markers for autosomal chromosomes and PAR1, which is present on the X and Y chromosomes. Mosaic LOY leads to an imbalance between parental alleles at PAR1 and the MoChA algorithm was used to detect allelic imbalances and decreases in coverage at PAR1 caused by mLOY. Input data included heterozygous markers with a minor allele frequency greater or equal to 1% in those where both alleles had 5 or more supporting reads. Additional filters included thinning markers to include one marker per 1,000 base pair window and removing variants overlapping with germline copy number variants (CNVs) previously identified. The MoChA caller was run with the extra option “--LRR-weight 0.0 --bdev-LRR-BAF 6.0” to disable the LRR+BAF model. The resulting mCA calls were filtered by excluding 1) those with lod score less than 5, 2) those on chromosome X but with inferred sex “unknown”, 3) those with estimated relative coverage higher than 2.9, 4) those with BAF deviation larger than 0.16 and relative coverage higher than 2.5. Steps 3 and 4 are used to exclude putative germline duplications. Cell fraction (CF) estimates for mCAs i.e. the proportion of cells in a sample with an mCA, were estimated using the MoChA algorithm. A threshold of 10% CF was used for defining low and high CF mLOY.

### Down-sampling

Detection of mLOY is based on allele specific depth at heterozygous sites in PAR1. Therefore, sensitivity for detection of mLOY can theoretically be impacted by the heterozygosity frequency, which can vary across individuals from different genetic similarity groups. To address this possible source of bias in the detection of mLOY, we down-sampled heterozygous sites in the AA, HA, and EA groups to match the distribution of the EAS group which has the lowest heterozygosity frequency as described in Jakubek et al.^15^. We have used this approach in previous work contrasting autosomal and chromosome X mCAs across ancestry groups^15^. After down-sampling we reassessed differences in mLOY frequency across groups.

### Estimation of genetic ancestry proportions

Ancestry estimates were derived using RFMix^54^. The reference panel consisted of 92 Europeans, 92 Africans from the 1000 Genomes project ^55^ and 92 Native American samples from HGDP^56^. SNPs with MAF>0.05 were used for these analyses and each chromosome was analyzed separately.

### SCAVENGE Analysis

Fine mapping was conducted using 248 index SNPs from mLOY association analyses conducted by Kessler et a ^16^. Using these index SNPs, we derived the fine-mapping posterior probabilities from the TOPMed mLOY association results following the statistical fine-mapping approach outlined in Chen et al^57^. To discover the mLOY relevant cell state at a high resolution, we used the SCAVENGE approach^31^ that discerns cells with the highest phenotypic enrichment through co-localization and investigates transitive connections through network propagation within the cell-to-cell network constructed by single-cell ATAC-seq data. A large hematopoiesis scATAC-seq dataset^58^ consisting of 33,819 cells across 23 cell types spanning from stem cells to their differentiated progeny was investigated. For dissecting the heterogeneity of mLOY relevant enrichment in HSCs and other cell types, we performed permutation testing 1,000 times to determine an empirical SCAVENGE TRS distribution for each cell to enable the binary classification of a cell that is enriched or depleted for mLOY. The number and proportion of these two cell states were examined across each cell type. The chromVAR was used to determine transcription factors (TFs) that likely drive cell-state-specific regulatory programs as previously described. The differences of TF motif activation scores among mLOY-relevant cell states in HSCs were examined using the Wilcoxon tests and the Bonferroni-adjusted P values were calculated to control for Type I errors.

### Circulating Blood Cell Counts

In TOPMed, the time interval between measurement of blood cell counts (CBC) and the blood draw used for mLOY determination varies widely^59^, ranging from ∼36 years prior to mLOY blood draw to 22 years after mLOY blood draw with a mean of -1 year (i.e., 1 year prior) and median of 0 years (**Figure S3**). Because we were mainly interested in whether mLOY predicts blood cell phenotypes (rather than vice versa), we elected to exclude from analysis any CBC measurement performed more than 3 years prior to the TOPMed blood draw. This removed about ⅓ of the data. Red blood cell, white blood cell and platelet quantitative traits were measured from freshly collected whole blood samples. In studies where multiple blood cell measurements were available, we selected a single measurement for each trait and each participant. Each trait was defined as follows: Hematocrit (HCT) as the percentage of volume of blood that is composed of red blood cells; Hemoglobin (HGB) as the mass per volume (grams per deciliter) of hemoglobin in the blood; Mean corpuscular hemoglobin (MCH) as the average mass in picograms of hemoglobin per red blood cell; Mean corpuscular hemoglobin concentration (MCHC) as the average mass concentration (grams per deciliter) of hemoglobin per red blood cell; Mean corpuscular volume (MCV) as the average volume of red blood cells, measured in femtoliters (fL); RBC count as the count of red blood cells in the blood, by number concentration in millions per microliter; Red cell distribution width (RDW) as the measurement of the ratio of variation in width to the mean width of the red blood cell volume distribution curve taken at +/- one CV; Total white blood cell count (WBC), neutrophil, monocyte, lymphocyte, eosinophil, basophil and platelet count were defined with respect to cell concentration in blood, measured in thousands/microliter. Mean platelet volume (MPV) was measured in fL.

### Calling CHIP, mCA, and Telomere Length

Somatic CHIP mutations were based on the TOPMed WGS data and as described in Bick et al.^30^ Telomere length was called using the TelSeq^60^ methodology from the TOPMed WGS data and detailed in Taub et al.^61^ Mosaic chromosomal alterations were also called using the WGS data as described in Jabubek et al.^15^

### Statistical Analyses- genetic ancestry groups

Association tests for mLOY and ancestry were run using the logistf package in R. The dependent variable was mLOY which was modeled as a binary trait. All analyses were adjusted for age, age^2^, and study phase. For some models smoking was also included and was modeled as a binary trait (never or ever smoker). In the latter analyses, only a subset of individuals with available smoking data were included.

### Statistical Analyses- mCAs and CHIP

For the analyses of autosomal mCA subtypes (A-mCA, L-mCA, M-mCA, U-mCA) and mLOY, autosomal mCAs were classified following the approach used by Niroula et al.^25^ These analyses excluded those with multiple mCAs. As with the ancestry analyses, mLOY was the dependent variable and the analysis was adjusted for age, age^2^, study phase. When smoking was included it was modeled as a binary phenotype (ever/never).

### Statistical Analysis of Allelic Shift in PAR1

We also tested for non-random loss of alleles at heterozygous sites in PAR1 using the binomial distribution. For all individuals with mLOY and heterozygous at a specific SNP, we used the read depth to score a shift toward the reference or the alternate allele and a binomial test to determine if there was a statistically significant deviation from an equal i.e. 50% chance of a shift to either allele at that SNP. We adjusted for multiple testing using the Bonferroni method (n=number of heterozygous SNPs). We also randomly selected an equal number of individuals without mLOY as a control set and re-ran this analysis. No SNP demonstrated a statistically significant allelic shift in this control set.

### Genetic Association Analyses

Genetic association analyses were run using the Michigan Encore server (https://encore.sph.umich.edu). Covariates used in all analyses include age, age^2^, study, genetic ancestry groups (AA, EA, HA, EAS), and the first 10 genetic principal components. The fast logistic regression model with kinship adjustment (saige-bin) threshold for single variant tests was utilized for the single variant analyses. For the single variant association test, variants with minor allele frequency (MAF) greater than 0.01% were included. The Encore saige-0.39 pipeline and genotype calls from TOPMed - Freeze 8 (Feb 2019) (GRCh38) were used for this analysis. We utilized the adaptive burden test (SKAT-O) for gene burden analyses. Variants with MAF less than 5% MAF were included in the gene burden analyses. The Encore epacts-3.4.1 pipeline and genotype calls from TOPMed - Freeze 8 (Feb 2019) (GRCh38) were used for this analysis.

### Gene expression analysis

We tested for an association between mLOY and RNA expression in blood in JHS and MESA. RNA was derived from peripheral blood mononuclear cells in JHS and MESA.

No hard thresholds were set on individual QC metrics for each RNA-seq sample; a PCA-based approach was used to label samples as outliers or non-outliers, and outlier samples were excluded from the eQTL scan. For each cohort/tissue combination, DESeq2 size factor-based normalization of gene counts was performed^62^, as implemented in pyqtl^63^ function deseq2_normalized_counts.

We dropped genes for which fewer than 10% of samples had a normalized count of at least 10, filled zeros with a value equal to ½ of the minimum observed non-zero value for that gene, log10 transformed the matrix, and then performed PCA. A sample was labeled as an outlier in PC space if any of the following criteria were met: 1) The sample’s Mahalanobis distance, computed with the top 5 PCs, corresponded to a chi-square p-value < 0.001. 2) Along any of the top 10 PCs, the sample’s absolute deviation from the median, normalized by the median absolute deviation across all samples for that PC, was >= 5.

For each tissue, samples for the cis-eQTL scan were selected via the following procedure: 1) Exclude samples that were outliers in PC space. 2) Exclude related participants. 3) Exclude samples with unclear sex based on gene expression (inferred using TPM values for genes XIST (on chrX) and RPS4Y1 (on chrY)).

For JHS we derived gene expression as in Wen et al.^64^ and inverse normalized. For MESA we followed the procedure outlined in Orchard et al.^67^. Associations were found using glm from the base version R package stats. Covariates were age, age^2^, race or ethnicity (in MESA), 10 genetic ancestry PCs for MESA and 2 genetic ancestry PCs for JHS. We included 10 RNA peer factors^65^ for MESA and 2 RNA peer factors for JHS. We included ever smoking status as a covariate in these analyses.

JHS and MESA were analyzed separately. Genes for which < 20% of samples have a TPM value of > 0.1 were dropped. The results were then meta-analyzed using METAL^66^ for 556 expression IDs (chromosome Y genes) common between the cohorts. We excluded samples where age at DNA blood draw for WGS and age at blood draw for RNA-seq were not equivalent. **Supplementary Table S23** displays the number of samples used in these analyses along with mLOY and smoking status.

## Data Availability

Data for each participating study can be accessed through dbGaP with the corresponding accession numbers:

| TOPMed Accession # | TOPMed Project | Parent Study Name | TOPMed Phase | Omics Center | Omics Support |
| --- | --- | --- | --- | --- | --- |
| phs000956 | Amish | Amish | 1 | Broad Genomics | 3R01HL121007-01S1 |
| phs001211 | AFGen | ARIC AFGen | 1 | Broad Genomics | 3R01HL092577-06S1 |
| phs001211 | VTE | ARIC | 2 | Baylor | 3U54HG003273-12S2 / HHSN268201500015C |
| phs001644 | AFGen | BioMe AFGen | 2.4 | MGI | 3UM1HG008853-01S2 |
| phs001143 | BAGS | BAGS | 1 | Illumina | 3R01HL104608-04S1 |
| phs001644 | BioMe | BioMe | 3 | Baylor | HHSN268201600033I |
| phs001644 | BioMe | BioMe | 3 | MGI | HHSN268201600037I |
| phs001612 | CARDIA | CARDIA | 3 | Baylor | HHSN268201600033I |
| phs000954 | CFS | CFS | 3.5 | NWGC | HHSN268201600032I |
| phs000954 | CFS | CFS | 1 | NWGC | 3R01HL098433-05S1 |
| phs001368 | CHS | CHS | 3 | Baylor | HHSN268201600033I |
| phs001368 | VTE | CHS VTE | 2 | Baylor | 3U54HG003273-12S2 / HHSN268201500015C |
| phs000951 | COPD | COPDGene | 1 | NWGC | 3R01HL089856-08S1 |
| phs000951 | COPD | COPDGene | 2 | Broad Genomics | HHSN268201500014C |
| phs000951 | COPD | COPDGene | 2.5 | Broad Genomics | HHSN268201500014C |
| phs000974 | AFGen | FHS AFGen | 1 | Broad Genomics | 3R01HL092577-06S1 |
| phs000974 | FHS | FHS | 1 | Broad Genomics | 3U54HG003067-12S2 |
| phs001218 | AA_CAC | GeneSTAR AA_CAC | 2 | Broad Genomics | HHSN268201500014C |
| phs001218 | GeneSTAR | GeneSTAR | legacy | Illumina | R01HL112064 |
| phs001218 | GeneSTAR | GeneSTAR | 2 | Psomagen | 3R01HL112064-04S1 |
| phs001345 | GENOA | HyperGEN_GENOA | 2 | NWGC | 3R01HL055673-18S1 |
| phs001345 | GENOA AA_CAC | AA_CAC | 2 | Broad Genomics | HHSN268201500014C |
| phs001359 | GOLDN | GOLDN | 2 | NWGC | 3R01HL104135-04S1 |
| phs001395 | HCHS_SOL | HCHS_SOL | 3 | Baylor | HHSN268201600033I |
| phs001293 | HyperGEN | HyperGEN_GENOA | 2 | NWGC | 3R01HL055673-18S1 |
| phs000964 | JHS | JHS | 1 | NWGC | HHSN268201100037C |
| phs001416 | AA_CAC | MESA AA_CAC | 2 | Broad Genomics | HHSN268201500014C |
| phs001416 | MESA | MESA | 2 | Broad Genomics | 3U54HG003067-13S1 |
| phs001032 | VU_AF | AFGen | 2 | Broad Genomics | 3R01HL092577-06S1 |

## Code Availability

Code to implement the LoY calling is available on GitHub https://github.com/auerlab.

## Acknowledgements

Molecular data for the Trans-Omics in Precision Medicine (TOPMed) program was supported by the National Heart, Lung and Blood Institute (NHLBI). The views expressed in this manuscript are those of the authors and do not necessarily represent the views of the National Heart, Lung, and Blood Institute; the National Institutes of Health; or the U.S. Department of Health and Human Services. See Supplementary Materials for extended acknowledgements.

## Author Contributions

P.L.A., P.S., A.P.R., and Y.A.J. conceived the study. P.L.A. supervised the work. A.P.R., P.S., D.W.F., L.M.R., Y.L., and V.G.S. assisted in directing the overall analyses. Y.A.J., A.S., A.P.R., J.C., L.M.R., B.T., and P.L.A. performed the statistical analyses. Y.A.J., J.B., X.M., and P.L.A. developed and implemented the bioinformatics and computational pipelines. K.B., B.M., E.B., R.J.F.L., M.F., M.C., D.D., J.B., B.P., N.H.C., A.D., R.M., K.N., L.R., M.M., S.R., P.P., J.S., R.I., B.H., D.A., T.H., J.R., S.S.R., J.C.M., and N.C. contributed to the design and conduct of the contributing TOPMed studies. Y.A.J. and P.L.A. drafted the manuscript. All authors reviewed and approved the paper.

