## Supplementary Methods for "Genomic and phenotypic correlates of mosaic loss of chromosome Y in blood"

**Participating TOPMed studies**

*Amish*

The Amish Complex Disease Research Program includes a set of large community-based studies focused largely on cardiometabolic health carried out in the Old Order Amish (OOA) community of Lancaster, Pennsylvania (http://medschool.umaryland.edu/endocrinology/amish/research-program.asp). The OOA population of Lancaster County, PA immigrated to the Colonies from Western Europe in the early 1700’s. There are now over 30,000 OOA individuals in the Lancaster area, nearly all of whom can trace their ancestry back 12-14 generations to approximately 700 founders. Investigators at the University of Maryland School of Medicine have been studying the genetic determinants of cardiometabolic health in this population since 1993. To date, over 7,000 Amish adults have participated in one or more of our studies.

Due to their ancestral history, the OOA are enriched for rare exonic variants that arose in the population from a single founder (or small number of founders) and propagated through genetic drift. Many of these variants have large effect sizes and identifying them can lead to new biological insights about health and disease. The parent study for this WGS project provides one (of multiple) examples. In our parent study, we identified through a genome-wide association analysis a haplotype that was highly enriched in the OOA that is associated with very high LDL-cholesterol levels. At the present time, the identity of the causative SNP – and even the implicated gene – is not known because the associated haplotype contains numerous genes, none of which are obvious lipid candidate genes. A major goal of the WGS that will be obtained through the NHLBI TOPMed Consortium will be to identify functional variants that underlie some of the large effect associations observed in this unique population.

*ARIC*

The ARIC study is a population-based prospective cohort study of cardiovascular disease sponsored by the National Heart, Lung, and Blood Institute (NHLBI). ARIC included 15,792 individuals, predominantly European American and African American, aged 45-64 years at baseline (1987-89), chosen by probability sampling from four US communities. Cohort members completed three additional triennial follow-up examinations, a fifth exam in 2011-2013, a sixth exam in 2016-2017, a seventh exam in 2018-2019, an eight exam in 2020, a ninth exam in 2021-2022, and a tenth exam in 2023. The ARIC study has been described in detail previously (PMC8667593; Wright JD, Folsom AR, Coresh J, et al. The ARIC (Atherosclerosis Risk In Communities) Study: JACC Focus Seminar 3/8. J Am Coll Cardiol. 2021 Jun 15;77(23):2939-2959).

*BAGS*

Epidemiologic studies of asthma have been underway in Barbados since 1991, when PI Barnes reported a relationship between modernization of the domestic environment in Barbados and increased risk of asthma. The baseline prevalence of asthma in Barbados is high (~20%), and from admixture analyses, we have determined that the proportion of African ancestry among Barbadian founders is similar to U.S. African Americans, rendering this a unique population to disentangle the genetic basis for asthma disparities among African ancestry populations in general. The primary outcome measure is asthma, and the approach for characterizing asthma in the Barbados population is based on the validated Respiratory Health Questionnaire (RHQ) designed from the 1978 American Thoracic Society questionnaire. Additional phenotype data include lung function measures, asthma severity, total serum IgE, and serum levels of various cytokines. In 1993, the *Barbados Asthma Genetics Study* (BAGS) was initiated on nuclear and extended asthmatic families who self-reported as African Caribbean, resulting in the first evidence for linkage for asthma and tIgE in an African-ancestry population, and the development of novel family-based methods. Recruitment into the BAGS program was enhanced through its involvement in the international *Genetics of Asthma International Network* (1999-2001) and the current sample of >1300 participants continues to grow through the efforts of collaborators and nursing staff at the Chronic Disease Research Centre in Barbados. Pediatric probands were recruited through referrals at local polyclinics or the Accident and Emergency Department at the Queen Elizabeth Hospital, and their nuclear and extended family members were subsequently recruited. All subjects gave verbal and written consent as approved by the Johns Hopkins Institutional Review Board (IRB) and the Barbados Ministry of Health.

In 2007 we performed a genome-wide association study (GWAS) on 655,352 SNPs using the Illumina Infinium™ II HumanHap650Y BeadChip v.1.0 (Illumina Inc.) on a subset of 1,000 Barbados participants. This represented the first GWAS of asthma focusing exclusively on populations of African ancestry, and data from this study also contributed to the NHLBI-supported EVE Consortium. BAGS also contributed 96 samples to Phase 2 of the Thousand Genomes Project (TGP). Subsequently, BAGS samples were included in the NHLBI-supported parent grant, entitled *New Approaches for Empowering Studies of Asthma in Populations of African Descent*“ (R01 HL104608-01), in which whole genome sequencing (WGS) was performed on ~1,000 individuals from North, Central, and South American and Caribbean and two West African populations. These populations constitute the *Consortium on Asthma among African-ancestry Populations in the Americas* (CAAPA), which aims to discover genes influencing risk for asthma, and catalog genetic diversity in descendants of the African Diaspora in the Americas. So far, CAAPA sequencing has greatly expanded the lexicon of human diversity, as we have observed >20% more variants than reported in the 1000 Genome Project (TGP). Using these WGS data, a custom, gene-centric SNP genotyping array was developed by Illumina, Inc., called the *African Diaspora Power Chip* (ADPC), to complement current, commercially available genome-wide chips, which provide sub-optimal tagging of genes among individuals of African ancestry. This ADPC was recently genotyped on all BAGS samples, with a goal of combining ADPC data with existing GWAS data from the 650Y to test for association with asthma. The initial goals of the parent grant did not include validating the ADPC. Moreover, the ADPC, combined with existing GWAS data, will be limited in detecting contributions of rare and structural variants, which may account for some of the “missing heritability” of asthma. We therefore are performing WGS on 1,100 asthmatics and family members from the BAGS, in order to (i) expand the CAAPA WGS dataset and thereby the genomic catalog of African ancestry for the research community; (ii) validate the ADPC by capturing information from both common and rare variants; and (iii) generate additional discovery of rare and structural variants that may control risk to asthma. Tools resulting from this study will result in substantial advancements in the technology available for identifying genes relevant to disease in under-represented minorities.

Given the data available on this large, deeply genotyped cohort from a relatively homogeneous environment representing an underrepresented minority group suffering most from asthma, the BAGS sample provides a unique opportunity to employ novel genomics.

*BioMe*

The Charles Bronfman Institute for Personalized Medicine at Mount Sinai Medical Center (MSMC), BioMe Biobank, founded in September 2007, is an ongoing, broadly-consented electronic health record-linked clinical care biobank that enrolls participants non-selectively from the Mount Sinai Medical Center patient population. The MSMC serves diverse local communities of upper Manhattan, including Central Harlem (86% African American), East Harlem (88% Hispanic/Latino), and Upper East Side (88% Caucasian/White) with broad health disparities.

*CARDIA*

The Coronary Artery Risk Development in Young Adults (CARDIA) Study is a study examining the development and determinants of clinical and subclinical cardiovascular disease and their risk factors. It began in 1985-1986 with a group of 5,115 black and white men and women aged 18-30 years. The participants were selected so that there would be approximately the same number of people in subgroups of race, gender, education (high school or less and more than high school) and age (18-24 and 25-30) in each of 4 centers: Birmingham, AL; Chicago, IL; Minneapolis, MN; and Oakland, CA.

*CFS*

The CFS is a genetic epidemiological study of 352 rigorously phenotyped families ascertained through probands with OSA identified through Cleveland, OH area sleep centers, neighborhood controls, and the spouses and first and second degree relatives of probands. Participants were studied on up to 4 exams between 1990-2006 with overnight sleep studies, standardized anthropometry; questionnaires; blood pressure; and spirometry. Fasting serum and ECGs are available from the last exam. Participants have a mean age of 37.7 years (African Americans) and 41.4 years (European Americans). Slightly more than 50% of the sample is female and 31% have moderate to severe OSA; 12.6% have diabetes, and 34.0% have hypertension. Asthma is reported in 19% and 13% of the African Americans and European Americans, respectively. Heritability analysis of traditional OSA traits as well as novel traits such as hypopnea duration (a marker of respiratory arousability) as well as overnight oxygenation (a marker of susceptibility to hypoxemia occurring with recurrent apneas) has shown that the latter traits are more heritable (h2 > 0.50) than traditional measures. Linkage analysis has identified peaks (and individual families contributing to peaks) for these traits. Through the Life After Linkage initiative (5R01HL113338), we further have aggregated and analyzed data on 19,798 individuals from 7 cohorts (Cleveland Family Study [CFS] plus ARIC, FHS, HCHS/SOL, MESA, MrOS, and Starr County) and conducted the largest GWAS to date of OSA traits.

*CHS*

The Cardiovascular Health Study (CHS) is a population-based cohort study of risk factors for coronary heart disease and stroke in adults 65 years and older conducted across four field centers 2. The original predominantly European ancestry cohort of 5,201 persons was recruited in 1989-1990 from random samples of people on Medicare eligibility lists from four US communities. Subsequently, an additional predominantly African-American cohort of 687 persons was enrolled for a total sample of 5,888. Institutional review committees at each field center approved the CHS, and participants gave informed consent. Blood samples were drawn from all participants at their baseline examination, and DNA was subsequently extracted from available samples. These analyses were limited to participants with available DNA who also consented to genetic studies. Participants were examined annually from enrollment to 1999 and continued to be under surveillance for stroke following 1999.

*COPDGene*

COPDGene (also known as the Genetic Epidemiology of COPD Study) is an NIH-funded, multicenter study. A study population of more than 10,000 smokers (1/3 African American and 2/3 non-Hispanic White) has been characterized with a study protocol including pulmonary function tests, chest CT scans, six minute walk testing, and multiple questionnaires. Five years after this initial visit, all available study participants are being brought back for a follow-up visit with a similar study protocol. This study has been used for epidemiologic and genetic studies. Previous genetic analysis in this study has been based on genome-wide SNP genotyping data. Approximately 1,900 subjects underwent whole genome sequencing in this NHLBI WGS project, including severe COPD subjects and resistant smoking controls. The COPDGene Study web site is: http://www.copdgene.org/.

*FHS*

FHS is a three-generation, single-site, community-based, ongoing cohort study that was initiated in 1948 to investigate prospectively the risk factors for CVD including stroke. It now comprises 3 generations of participants: the Original cohort followed since 1948; their Offspring and spouses of the Offspring, followed since 1971 4; and children from the largest Offspring families enrolled in 2002 (Gen 3). The Original cohort enrolled 5,209 men and women who comprised two-thirds of the adult population then residing in Framingham, MA. Survivors continue to receive biennial examinations. The Offspring cohort comprises 5,124 persons (including 3,514 biological offspring) who have been examined approximately once every 4 years. The Gen 3 cohort contains 4,095 participants.

*GeneSTAR*

In 1982 The Johns Hopkins Sibling and Family Heart Study was created to study patterns of coronary heart disease and related risk factors in families with early-onset coronary disease, identified from10 Baltimore area hospitals. Renamed in 2003, the Genetic Study of Atherosclerosis Risk (GeneSTAR) continues to study mechanisms of coronary heart disease and stroke in families using novel models and exciting new methods. GeneSTAR is a family-based study including initially healthy brothers and sisters identified from probands with early-onset coronary disease, along with the healthy offspring of the siblings and the probands. The goal is to discover and amplify mechanisms of stroke and coronary heart disease. Our African American and European American family cohort has undergone extensive screening, genetic testing, and follow-up for new cardiovascular disease, stroke, and other clinical events for 5 to 38 years.

*HCHS/SOL*

The Hispanic Community Health Study/Study of Latinos (HCHS/SOL) is a multi-center study of Hispanic/Latino populations with the goal of determining the role of acculturation in the prevalence and development of diseases, and to identify other traits that impact Hispanic/Latino health. The study is sponsored by the National Heart, Lung, and Blood Institute (NHLBI) and other institutes, centers, and offices of the National Institutes of Health (NIH). Recruitment began in 2006 with a target population of 16,000 persons of Cuban, Puerto Rican, Dominican, Mexican or Central/South American origin. Participants were recruited through four sites affiliated with San Diego State University, Northwestern University in Chicago, Albert Einstein College of Medicine in Bronx, New York, and the University of Miami. Recruitment was implemented through a two-stage area household probability design. The study enrolled 16,415 participants who were self-identified Hispanic/Latino and aged 18-74 years, and the extensive psycho-social and clinical assessments were conducted during 2008-2011. Annual telephone follow-up interviews are ongoing since study inception. During the 2014-2017 second visit, the participants were re-examined again for various health outcomes of interest.

*GENOA*

Details of the TOPMed Project: Hypertension Genetic Epidemiology Network and Genetic Epidemiology Network of Arteriopathy study can be found at: https://topmed.nhlbi.nih.gov/

*GOLDN*

Details of the TOPMed Project: Genetics of Lipid Lowering Drugs and Diet Network project can be found at: https://topmed.nhlbi.nih.gov/

*HyperGEN*

Details of the TOPMed Project: Hypertension Genetic Epidemiology Network and Genetic Epidemiology Network of Arteriopathy study can be found at: https://topmed.nhlbi.nih.gov/

*JHS*

The Jackson Heart Study (JHS, https://www.jacksonheartstudy.org/jhsinfo/) is a large, community-based, observational study whose participants were recruited from urban and rural areas of the three counties (Hinds, Madison and Rankin) that make up the Jackson, MS metropolitan statistical area (MSA). Participants were enrolled from each of 4 recruitment pools: random, 17%; volunteer, 30%; currently enrolled in the Atherosclerosis Risk in Communities (ARIC) Study, 31% and secondary family members, 22%. Recruitment was limited to non-institutionalized adult African Americans 35-84 years old, except in a nested family cohort where those 21 to 34 years of age were also eligible. The final cohort of 5,306 participants included 6.59% of all African American Jackson MSA residents aged 35-84 during the baseline exam (N-76,426, US Census 2000). Among these, approximately 3,700 gave consent that allows genetic research and deposition of data into dbGaP, with 3406 participants with post-QC TOPMed whole genome sequencing data. Major components of three clinic examinations (Exam 1 – 2000-2004; Exam 2 – 2005-2008; Exam 3 – 2009-2013) include medical history, physical examination, blood/urine analytes and interview questions on areas such as: physical activity; stress, coping and spirituality; racism and discrimination; socioeconomic position; and access to health care. Extensive clinical phenotyping includes anthropometrics, electrocardiography, carotid ultrasound, ankle-brachial blood pressure index, echocardiography, CT chest and abdomen for coronary and aortic calcification, liver fat, and subcutaneous and visceral fat measurement, and cardiac MRI. At 12-month intervals after the baseline clinic visit (Exam 1), participants have been contacted by telephone to: update information; confirm vital statistics; document interim medical events, hospitalizations, and functional status; and obtain additional sociocultural information. Questions about medical events, symptoms of cardiovascular disease and functional status are repeated annually. Ongoing cohort surveillance includes abstraction of medical records and death certificates for relevant International Classification of Diseases (ICD) codes and adjudication of nonfatal events and deaths. CMS data are currently being incorporated into the dataset.

*MESA*

The MESA study is a study of the characteristics of subclinical cardiovascular disease (disease detected non-invasively before it has produced clinical signs and symptoms) and the risk factors that predict progression to clinically overt cardiovascular disease or progression of the subclinical disease 7. MESA researchers study a diverse, population-based sample of 6,814 asymptomatic men and women aged 45-84. Thirty-eight percent of the recruited participants are White, 28 percent African-American, 22 percent Hispanic, and 12 percent Asian, predominantly of Chinese descent. Participants were recruited from six field centers across the United States: Wake Forest University, Columbia University, Johns Hopkins University, University of Minnesota, Northwestern University and the University of California - Los Angeles.

*VU_AF*

The Vanderbilt Atrial Fibrillation (AF) Registry was founded in 2001. Patients with AF and family members are prospectively enrolled. At enrollment a detailed past medical history is obtained along with an AF symptom severity assessment. Blood samples are obtained for DNA extraction. Patients are followed longitudinally along with serial collection of AF symptom severity assessments.

**Supplementary Methods**

**Comparison of LoY calls from TOPMed WGS data and array-based calls**

We compared LoY calls from the TOPMed WGS data to array-based calls from the MESA study. There were 2,450 males in MESA typed on the Affymetrix 6.0 SNP array. In MESA, all samples for the WGS and array-based genotyping were collected at the same time. To call LoY with the array data in MESA, we only considered heterozygous markers in the PAR1 region of chrX. In order to make fair comparisons between the WGS and array data, we used the haplotypes from the TOPMed WGS data and substituted the allelic depth from the WGS data with the BAF and LRR from the array. Similar to the procedure we used for calling LoY with the WGS data, the MoChA caller was run with the extra option “--LRR-weight 0.0 --bdev-LRR-BAF 6.0” to disable the LRR+BAF mode. We also used the flag “regression="--adjust-BAF-LRR -1 --regress-BAF-LRR -1" to avoid batch effects when processing array data.

Subsequent calls were then excluded similarly to WGS: 1) those with LOD scores less than 5, 2) those with estimated relative coverage higher than 2.9, and 3) those with BAF deviation larger than 0.16 and relative coverage higher than 2.5. Only mCA events that were classified as “loss” in the PAR1 region were considered to be LoY.

Of the 281 LoY events that were identified via WGS in the 2,450 males in MESA, 54 (19%) were also identified as LoY based on the array. Of these 281 LoY events, 174 were determined to be high clonal fraction (CF>10%) events and 107 were determined to be low CF (CF<=10%) events. The array data successfully identified 54 (31%) of the high CF events and none of the low CF events. In total the arrays identified 54 total LoY events, all of which were also called using the WGS data.

In summary, the array-based LoY calls provide validation of the WGS-based callset, both by directly comparing concordance of LoY calls as well as by analysis of the LRRs on chrY. Of note, the lower CF LoY events were entirely missed by the arrays suggesting that WGS is more sensitive for detecting low CF LoY events. The main difference between the WGS and array-based calls appears to be the density of markers in PAR1. In the MESA samples, the median number of heterozygous markers in PAR1 was 93 in the array and 1,022 in the WGS. As has been previously reported, mCA detection is highly influenced by marker density with higher density data providing more power to detect mCAs. This is likely the primary driver of the increased sensitivity of WGS-based data to detect LoY compared to the Affymetrix 6.0 SNP array.

As a final validation step, we considered normalized read depth on chromosome Y as an indicator of LoY. Stratifying by the TOPMed sequencing center that processed and sequenced the samples, we found that the WGS-based LoY calls tracked with normalized chromosome Y read depth (**Figure S5**). The distribution of normalized chrY read depth was distinctly different for high CF events, whereas the normalized chrY read depth was not as distinguishable for low CF events (**Figure S6**). This suggests that haplotype-based callers, such as MoCha, are essential for identifying LoY events across a range of clone sizes.

**Supplementary Figures**

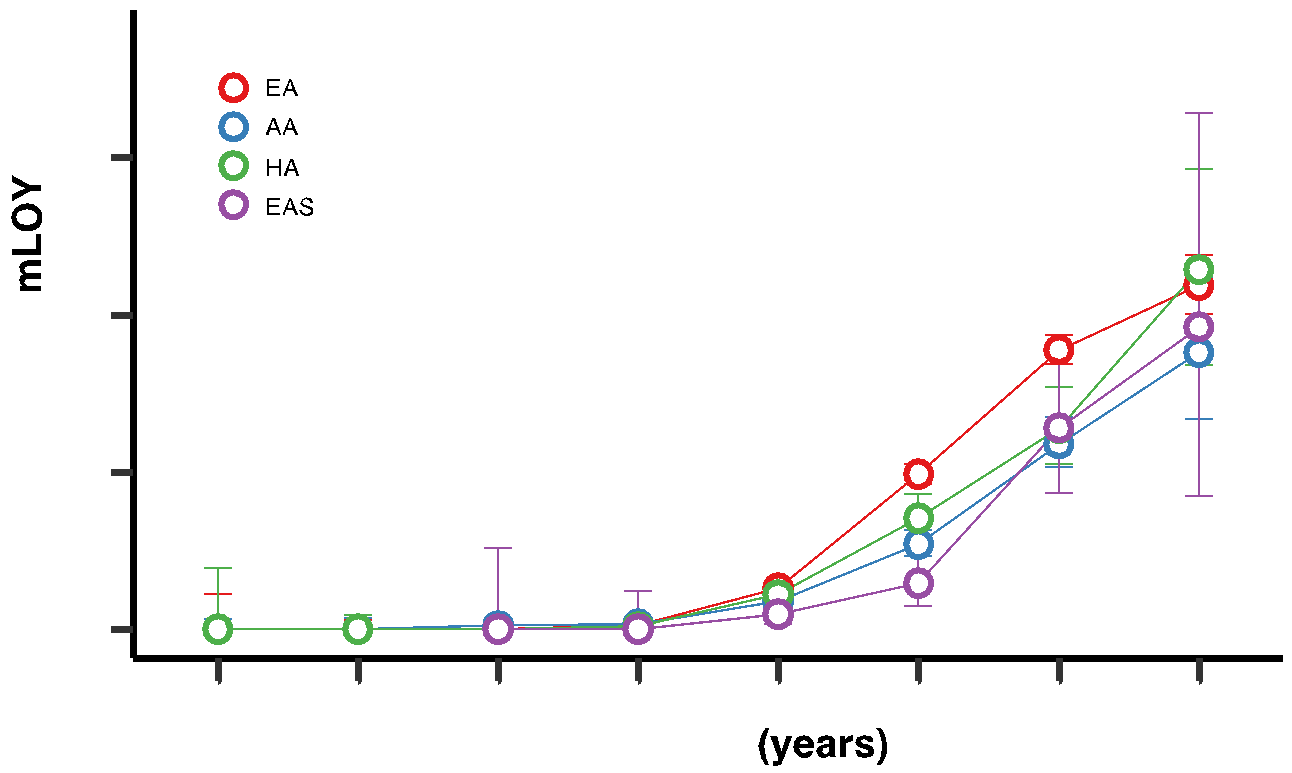

**Figure S1:** Fraction of individuals with mosaic loss of chromosome Y (mLOY) across age bins.

**A**

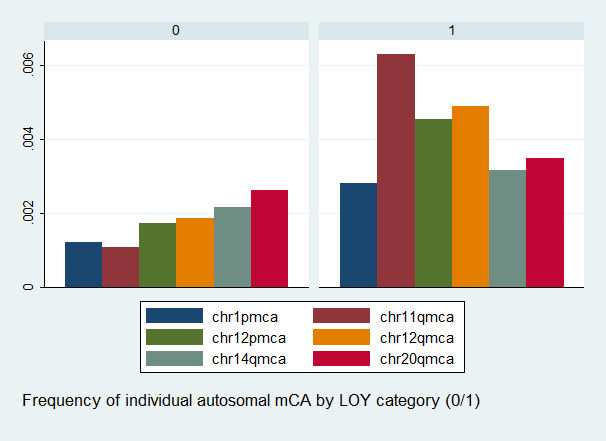

**B**

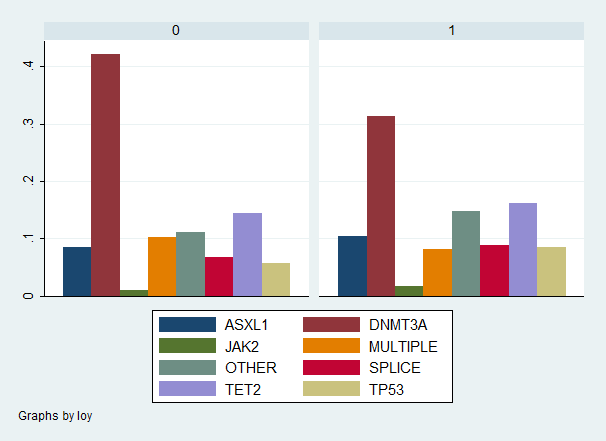

**Figure S2:** **A)** Frequency of individual 6 common autosomal mosaic chromosomal alterations by mosaic loss of chromosome Y (mLOY) status (0 = no mLOY; 1 = with mLOY). **B)** Frequency of individuals with mutations in CHIP genes by mLOY status.

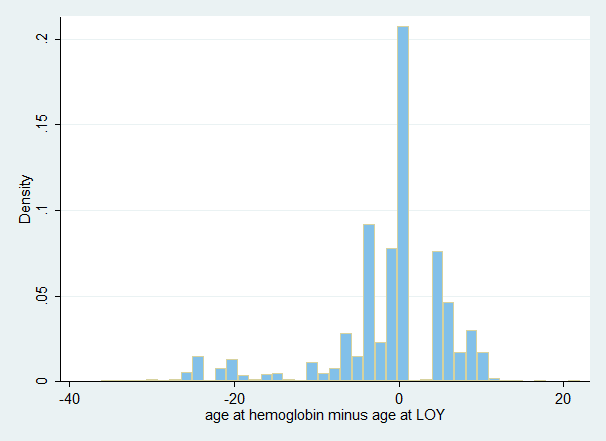

**Figure S3:** Distribution of age at blood draw (circulating blood-cell counts) minus age at DNA capture for mLOY determination.

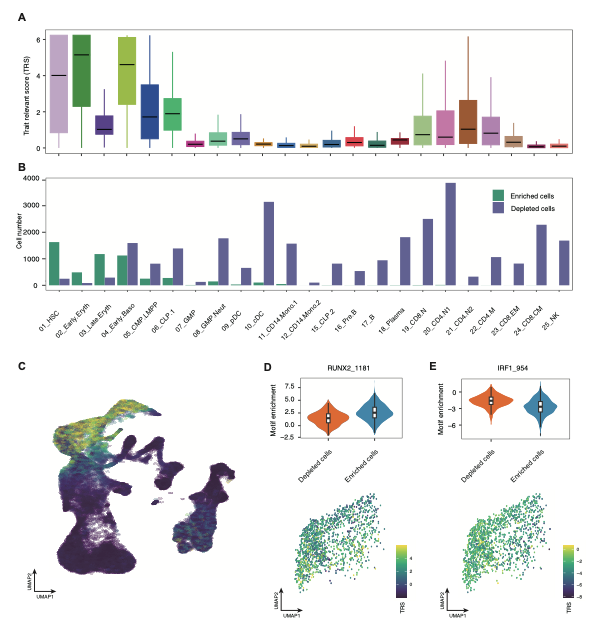

**Figure S4: A)** Mosaic loss of Y (mLOY) trait relevant score (TRS) at the cell type level. **B)** The number of cells for each cell state (enriched or depleted) **C)** MLOY TRS at the single cell level projected onto uniform manifold approximation and projection (UMAP). The transcription factor motif enrichment scores for **D)** RUNX2_1181 and **E)** IRF1_954 at the cell state and single-cell level.

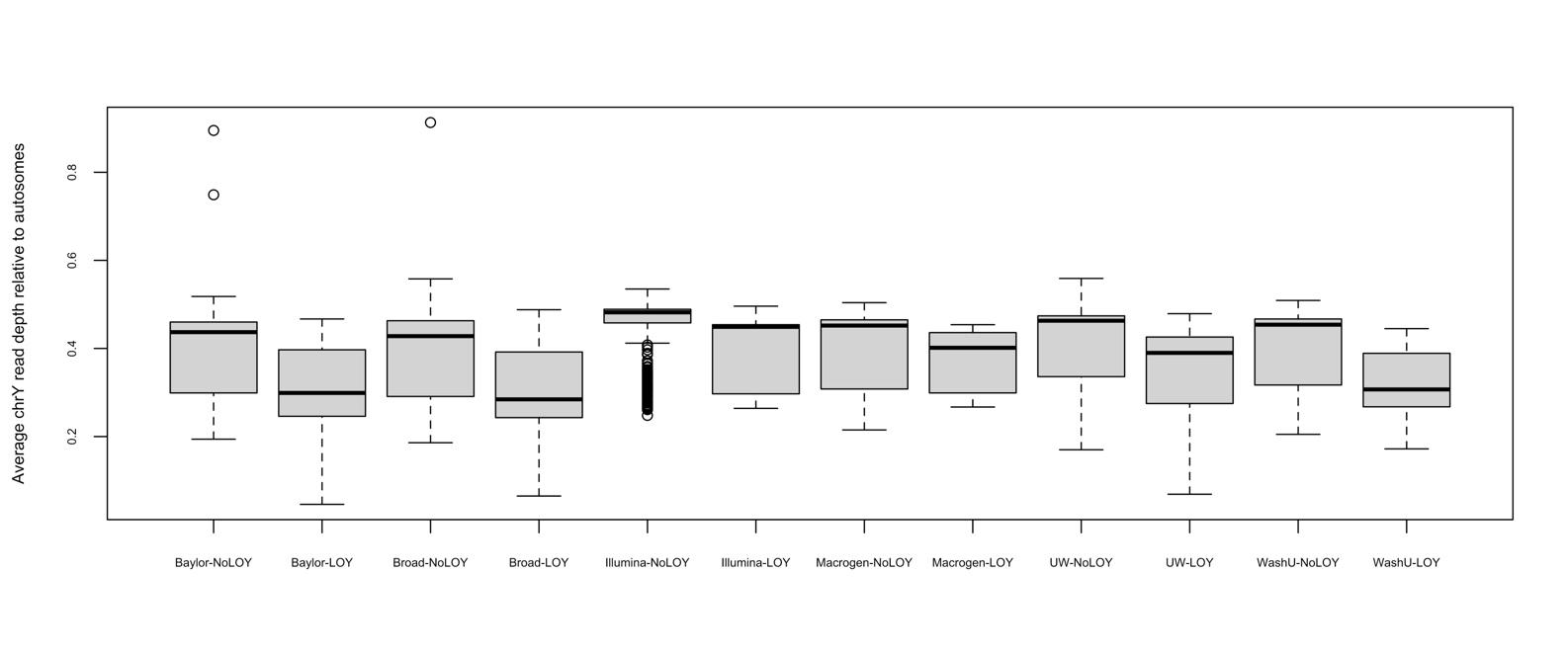

**Figure S5:** Distribution of chromosome Y (chrY) read depths relative to autosome for individuals with mosaic loss of Y (LOY) and those without (NoLOY) split by sequencing center.

**A**

High CF mLOY

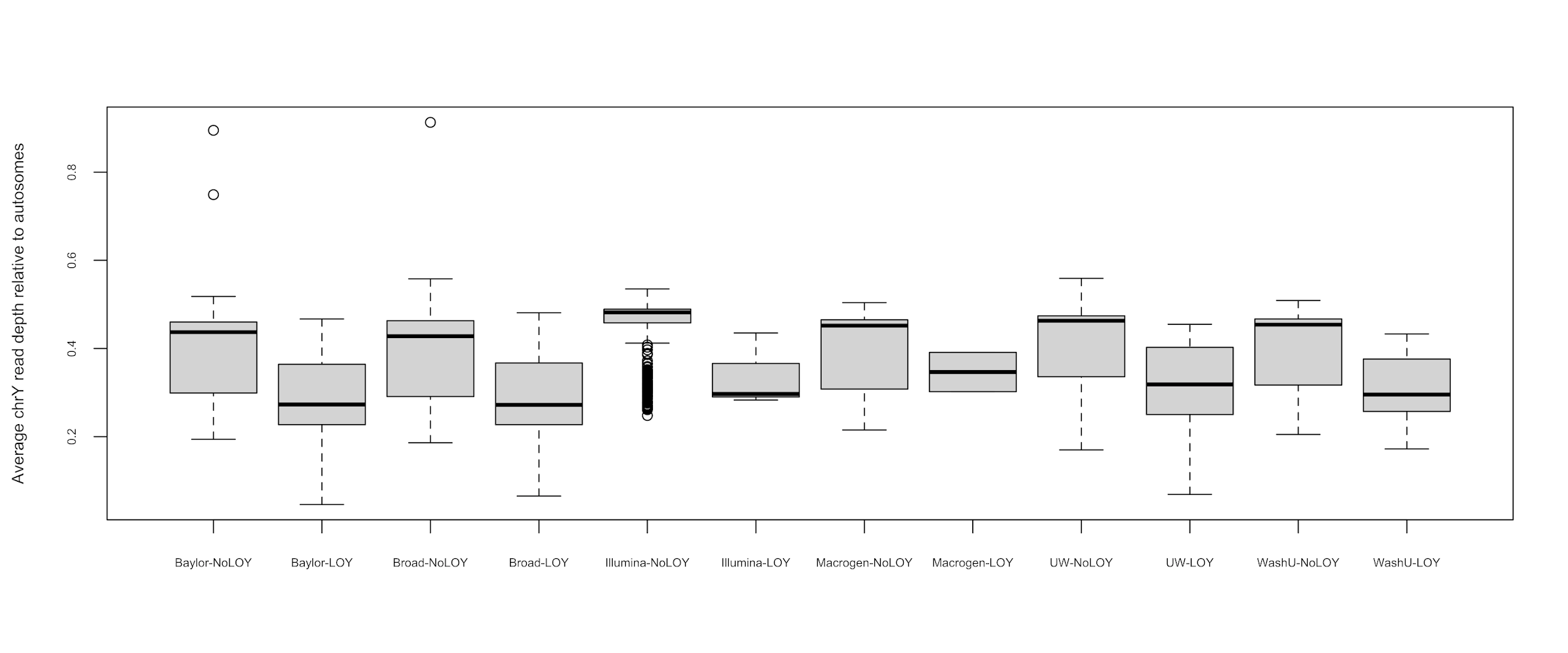

**B**

Low CF mLOY

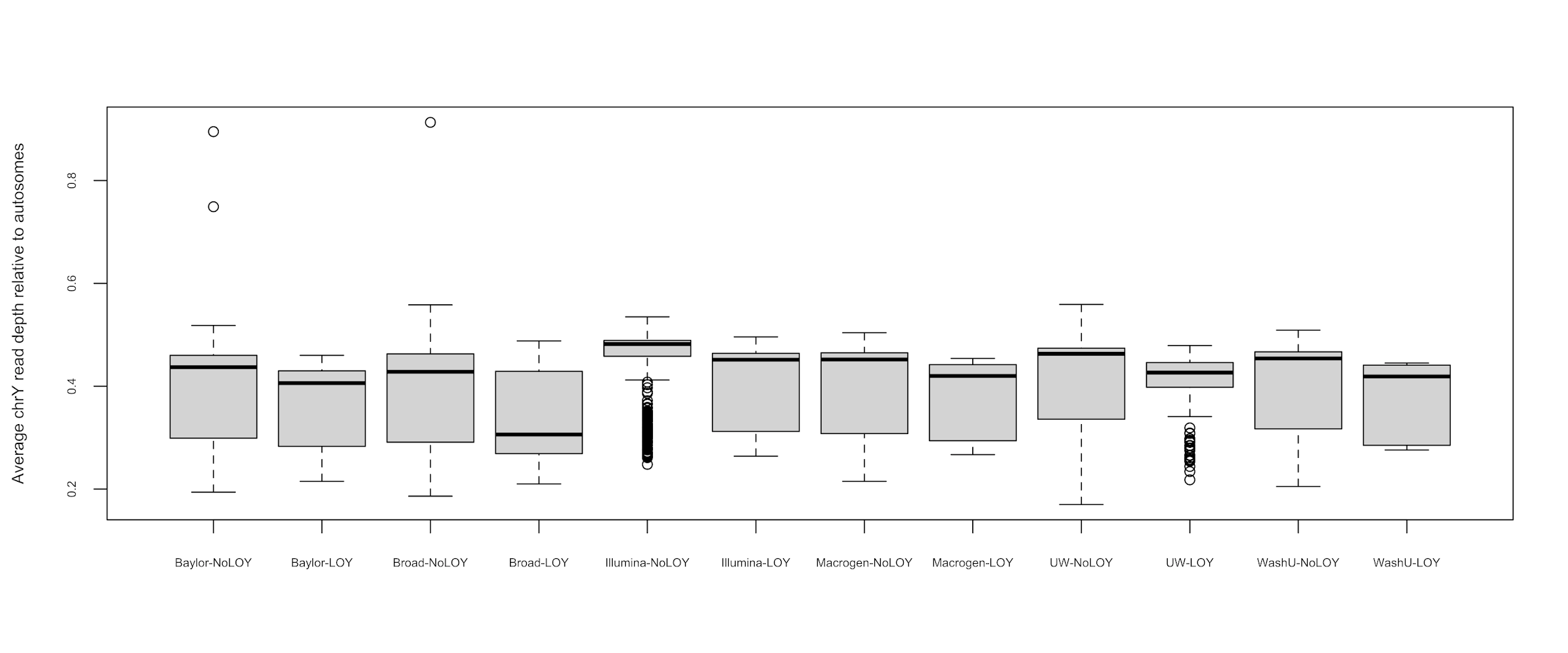

**Figure S6:** Distribution of chromosome Y (chrY) read depths relative to autosome for individuals with mosaic loss of Y (LOY) and those without (NoLOY) split by sequencing center. **A)** shows data for high cell fraction (CF) LOY (>= 10%) and **B)** for low CF LOY (< 10%).

**Extended Acknowledgements**

Core support including centralized genomic read mapping and genotype calling, along with variant quality metrics and filtering were provided by the TOPMed Informatics Research Center (3R01HL-117626-02S1; contract HHSN268201800002I). Core support including phenotype harmonization, data management, sample-identity QC, and general program coordination were provided by the TOPMed Data Coordinating Center (R01HL-120393; U01HL-120393; contract HHSN268201800001I).

Amish: The TOPMed component of the Amish Research Program was supported by NIH grants R01 HL121007, U01 HL072515, and R01 AG18728.

ARIC: The Atherosclerosis Risk in Communities study has been funded in whole or in part with Federal funds from the National Heart, Lung, and Blood Institute, National Institutes of Health, Department of Health and Human Services, under Contract nos. (75N92022D00001, 75N92022D00002, 75N92022D00003, 75N92022D00004, 75N92022D00005). The authors thank the staff and participants of the ARIC study for their important contributions.

BAGS: We gratefully acknowledge the contributions of Pissamai and Trevor Maul, Paul Levett, Anselm Hennis, P. Michele Lashley, Raana Naidu, Malcolm Howitt and Timothy Roach, and the numerous health care providers, and community clinics and co-investigators who assisted in the phenotyping and collection of DNA samples, and the families and patients for generously donating DNA samples to the Barbados Asthma Genetics Study (BAGS). Funding for BAGS was provided by National Institutes of Health (NIH) R01HL104608, R01HL087699, and HL104608 S1.

BioMe: The Mount Sinai BioMe Biobank has been supported by The Andrea and Charles Bronfman Philanthropies and in part by Federal funds from the NHLBI and NHGRI (U01HG00638001; U01HG007417; X01HL134588). We thank all participants in the Mount Sinai Biobank. We also thank all our recruiters who have assisted and continue to assist in data collection and management and are grateful for the computational resources and staff expertise provided by Scientific Computing at the Icahn School of Medicine at Mount Sinai.

CARDIA: The Coronary Artery Risk Development in Young Adults Study (CARDIA) is conducted and supported by the National Heart, Lung, and Blood Institute (NHLBI) in collaboration with the University of Alabama at Birmingham (HHSN268201800005I & HHSN268201800007I), Northwestern University (HHSN268201800003I), University of Minnesota (HHSN268201800006I), and Kaiser Foundation Research Institute (HHSN268201800004I). CARDIA was also partially supported by the Intramural Research Program of the National Institute on Aging (NIA) and an intra‐agency agreement between NIA and NHLBI (AG0005).

CFS: The Cleveland Family Study has been supported in part by National Institutes of Health grants [R01-HL046380, KL2-RR024990, R35-HL135818, and R01-HL113338].

CHS: This Cardiovascular Health Study research was supported by NHLBI contracts HHSN268201200036C, HHSN268200800007C, HHSN268201800001C, N01HC55222, N01HC85079, N01HC85080, N01HC85081, N01HC85082, N01HC85083, N01HC85086, 75N92021D00006; and NHLBI grants U01HL080295, R01HL087652, R01HL105756, R01HL103612, R01HL120393, and U01HL130114 with additional contribution from the National Institute of Neurological Disorders and Stroke (NINDS). Additional support was provided through R01AG023629 from the National Institute on Aging (NIA). A full list of principal CHS investigators and institutions can be found at CHS-NHLBI.org.

COPDGene: The COPDGene project described was supported by Award Number U01 HL089897 and Award Number U01 HL089856 from the National Heart, Lung, and Blood Institute. The content is solely the responsibility of the authors and does not necessarily represent the official views of the National Heart, Lung, and Blood Institute or the National Institutes of Health. COPDGene is also supported by the COPD Foundation through contributions made to an Industry Advisory Board that has included AstraZeneca, Bayer Pharmaceuticals, Boehringer-Ingelheim, Genentech, GlaxoSmithKline, Novartis, Pfizer, and Sunovion. A full listing of COPDGene investigators can be found at: <http://www.copdgene.org/directory>

FHS: The Framingham Heart Study (FHS) acknowledges the support of contracts NO1-HC-25195 and HHSN268201500001I from the National Heart, Lung and Blood Institute and grant supplement R01 HL092577-06S1 for this research. We also acknowledge the dedication of the FHS study participants without whom this research would not be possible.

GeneSTAR: GeneSTAR was supported by the National Institutes of Health/National Heart, Lung, and Blood Institute (U01 HL72518, HL087698, HL112064, HL11006, HL118356) and by a grant from the National Institutes of Health/National Center for Research Resources (M01-RR000052) to the Johns Hopkins General Clinical Research Center. We would like to thank our participants and staff for their valuable contributions.

GENOA: Support for GENOA was provided by the National Heart, Lung and Blood Institute (U01 HL054457, U01 HL054464, U01 HL054481, R01 HL119443, and R01 HL087660) of the National Institutes of Health.

GOLDN: GOLDN biospecimens, baseline phenotype data, and intervention phenotype data were collected with funding from National Heart, Lung and Blood Institute (NHLBI) grant U01 HL072524. Whole-genome sequencing in GOLDN was funded by NHLBI grant R01 HL104135 and supplement R01 HL104135-04S1.

HCHS/SOL: The Hispanic Community Health Study/Study of Latinos is a collaborative study supported by contracts from the National Heart, Lung, and Blood Institute (NHLBI) to the University of North Carolina (HHSN268201300001I / N01-HC-65233), University of Miami (HHSN268201300004I / N01-HC-65234), Albert Einstein College of Medicine (HHSN268201300002I / N01-HC-65235), University of Illinois at Chicago – HHSN268201300003I / N01-HC-65236 Northwestern Univ), and San Diego State University (HHSN268201300005I / N01-HC-65237). The following Institutes/Centers/Offices have contributed to the HCHS/SOL through a transfer of funds to the NHLBI: National Institute on Minority Health and Health Disparities, National Institute on Deafness and Other Communication Disorders, National Institute of Dental and Craniofacial Research, National Institute of Diabetes and Digestive and Kidney Diseases, National Institute of Neurological Disorders and Stroke, NIH Institution-Office of Dietary Supplements.

HyperGEN: The HyperGEN Study is part of the National Heart, Lung, and Blood Institute (NHLBI) Family Blood Pressure Program; collection of the data represented here was supported by grants U01 HL054472 (MN Lab), U01 HL054473 (DCC), U01 HL054495 (AL FC), and U01 HL054509 (NC FC). The HyperGEN: Genetics of Left Ventricular Hypertrophy Study was supported by NHLBI grant R01 HL055673 with whole-genome sequencing made possible by supplement -18S1.

JHS: The Jackson Heart Study (JHS) is supported and conducted in collaboration with Jackson State University (HHSN268201800013I), Tougaloo College (HHSN268201800014I), the Mississippi State Department of Health (HHSN268201800015I) and the University of Mississippi Medical Center (HHSN268201800010I, HHSN268201800011I and HHSN268201800012I) contracts from the National Heart, Lung, and Blood Institute (NHLBI) and the National Institute on Minority Health and Health Disparities (NIMHD). The authors also wish to thank the staff and participants of the JHS.

MESA: Whole genome sequencing (WGS) for the Trans-Omics in Precision Medicine (TOPMed) program was supported by the National Heart, Lung and Blood Institute (NHLBI). WGS for “NHLBI TOPMed: Multi-Ethnic Study of Atherosclerosis (MESA)” (phs001416.v1.p1) was performed at the Broad Institute of MIT and Harvard (3U54HG003067-13S1). Centralized read mapping and genotype calling, along with variant quality metrics and filtering were provided by the TOPMed Informatics Research Center (3R01HL-117626-02S1). Phenotype harmonization, data management, sample-identity QC, and general study coordination, were provided by the TOPMed Data Coordinating Center (3R01HL-120393-02S1), and TOPMed MESA Multi-Omics (HHSN2682015000031/HSN26800004). The MESA projects are conducted and supported by the National Heart, Lung, and Blood Institute (NHLBI) in collaboration with MESA investigators. Support for the Multi-Ethnic Study of Atherosclerosis (MESA) projects are conducted and supported by the National Heart, Lung, and Blood Institute (NHLBI) in collaboration with MESA investigators. Support for MESA is provided by contracts 75N92020D00001, HHSN268201500003I, N01-HC-95159, 75N92020D00005, N01-HC-95160, 75N92020D00002, N01-HC-95161, 75N92020D00003, N01-HC-95162, 75N92020D00006, N01-HC-95163, 75N92020D00004, N01-HC-95164, 75N92020D00007, N01-HC-95165, N01-HC-95166, N01-HC-95167, N01-HC-95168, N01-HC-95169, UL1-TR-000040, UL1-TR-001079, UL1-TR-001420, UL1TR001881, DK063491, and R01HL105756. The authors thank the other investigators, the staff, and the participants of the MESA study for their valuable contributions. A fill list of participating MESA investigators and institutes can be found at <http://www.mesa-nhlbi.org>.
